# Evaluation of a High-Acuity Next Day Clinic for Hospital Admission Avoidance: A 13-Month Cohort Study

**DOI:** 10.64898/2026.01.13.26344042

**Authors:** Richard K. Leuchter, Jaclyn Spiegel, William B. Turner, Paul Salama, Scott Lundberg, Mandi Occhiuto, Oleg Melamed, Vinhfield Ta, Vimol Reepolrujee, Anthony Simmons, Sitaram Vangala, Tristan Tibbe, Ben Waterman, Soma Wali

**Author notes:** **Address correspondence to**: Richard K. Leuchter, MD, Division of General Internal Medicine and Health Services Research, David Geffen School of Medicine at UCLA, 1100 Glendon Avenue, #726, Los Angeles, CA 90024. **Funding/support:** Dr. Leuchter was funded by NIH-NHLBI, grant no. 1K38 HL164955-01. The funders had no role in considering the study design or in the collection, analysis, interpretation of data, writing of the report, or decision to submit the article for publication. **Conflict of interest disclosures:** None of the authors disclose any conflict of interests including relevant financial interests, activities, relationships, and affiliations relevant to the subject of this manuscript.

## Abstract

**Importance:** Hospital capacity constraints and rising healthcare costs necessitate innovative models for delivering acute care. While various hospital-substitution models exist, challenges in scalability and long-term viability persist.

**Objective:** To evaluate the feasibility and safety of a novel, high-acuity Next Day Clinic (NDC) as an alternative to hospitalization for select acutely ill emergency department (ED) patients.

**Design, Setting, and Participants:** Retrospective matched cohort study of patients referred to the NDC between July 1, 2023-July 31, 2024, matched to patients seen in the ED during the year prior to NDC launch, within a large academic safety-net hospital.

**Intervention:** High-acuity outpatient therapy for one or more consecutive days in the NDC, consisting of daily IV antibiotics or diuretics, STAT labs, and rapid turnaround imaging and cardiodiagnostics.

**Main Outcomes and Measures:** Days alive and out of hospital (DAOH) in the 30 days following the index ED visit. Secondary outcomes were the number of hospital bed-days avoided, as well as 30- day ED revisits, hospital readmissions, and mortality.

**Results:** The NDC had 1009 encounters (mean age, 54.4 years [SD 14.6]; 448 female [44%]) during the study period, 420 (42%) of which were referred from the ED. Of these, 298 (71%) matched to 4666 ED visits (mean age, 53.3 years [SD 15.2]; 2019 female [43%]) in the year prior to NDC launch on age, sex, the first set of laboratory and vital sign data obtained in the ED (i.e., presenting illness severity), and an exact match on primary diagnosis group. Unadjusted mean DAOH in the NDC cohort was 29.5 days (SD 2.3) compared to 24.9 days (SD 5.5) in the control cohort. Adjusting for the same features in the matching algorithm showed NDC treatment was associated with an average of 3.85 (SD 0.20) more DAOH compared to hospitalization (p<0.001), translating to 358-1294 hospital bed-days saved over the study period. NDC patients had significantly higher rates of 30-day ED revisits per 100 encounters (20.5 versus 13.0, p<0.001), but significantly lower rates of 30-day hospital readmissions per 100 encounters (5.7 versus 11.0, p<0.001) and morality (0% versus 0.9%, p<0.001).

**Conclusions and Relevance:** The NDC is a feasible and safe alternative to hospitalization, and promising strategy for managing ED and hospital capacity and reducing healthcare expenditures.

**KEY POINTS:** *Question:* Is a high-acuity Next Day Clinic (NDC) a feasible and safe alternative to hospitalization for acutely ill emergency department (ED) patients?

*Findings:* In this matched cohort study of 1009 NDC encounters, 298 hospital admission avoidance referrals were matched with 4666 historical controls. Each avoided hospitalization through the NDC was associated with an average of 3.85 more days alive and out of the hospital over 30 days, lower readmissions and mortality, and a total of 358–1294 hospital bed-days saved.

*Meaning:* A centralized, high-acuity outpatient clinic may safely substitute for hospitalization, reducing hospital capacity strain and healthcare expenditures.

## INTRODUCTION

The U.S. is facing a critical hospital bed shortage as early as 2032 unless novel care models are developed to reduce the number of hospitalizations.^1^ To mitigate this potential crisis, the management of 10-20% of hospitalizations could be shifted from hospitals to office-based settings,^2–4^ but there are currently no widely disseminatable acute care models that can replace hospitalizations at-scale within the U.S.^5–7^

A spectrum of hospital admission avoidance strategies has emerged, ranging from remote patient monitoring programs to Hospital at Home.^8–11^ While these models have shown success, they face substantial logistical and implementation barriers preventing their widespread adoption.^5,12,13^ Hospital at Home, for example, faces startup costs on the order of $1.5-2 million,^14^ intensive ongoing human capital requirements,^15,16^ and do not enjoy the same economies of scale as centralized care models. Most concerningly, its reliance on a federal reimbursement waiver, which lapsed as of September 30, 2025 causing numerous Hospital at Home programs to fail, raises concerns about its long-term viability.^17^ There is an urgent need for acute care models that can centralize hospital-level services in a cost-effective outpatient setting, thereby improving health system resilience while simultaneously driving down the cost of care.

This study introduces and evaluates a novel hospital admission avoidance model: the Next Day Clinic (NDC). The NDC model delivers pre-planned, multi-day, rapid hospital-level services in a dedicated outpatient clinic, providing an alternative to inpatient admission for patients in the emergency department (ED). This study sought to evaluate the healthcare utilization metrics and safety of patients treated in the NDC, compared with a matched cohort of traditionally hospitalized patients.

## METHODS

### Study Design and Setting

We conducted a retrospective cohort study within Olive View-UCLA Medical Center (OVMC), a 355-bed, academic, county-affiliated public safety-net hospital in Los Angeles County. The study protocol was reviewed by the Olive View-UCLA Institutional Review Board, which issued a waiver of informed consent for this retrospective review of deidentified EHR data.

### Patient Population

#### Referral

All patients were referred to the NDC from either the ED or hospital wards from July 1, 2023 to July 31, 2024 (Figure 1). For referrals from the ED (called “hospital admission avoidance” referrals), the treating ED physician had to determine the patient as requiring hospital-level care for an acute medical condition, but deemed them clinically stable and logistically suitable for management in an intensive outpatient setting provided high-intensity treatment could be arranged. Referrals were reviewed for appropriateness by the on-call program attending who was either an ED physician or triage hospitalist, and patients who would have been discharged anyways from the ED were not accepted to the NDC. For referrals from the inpatient setting (called “early discharge” referrals), patients were required to be on a hospitalist service and need at least one more night in the hospital, but could be discharged early if intensive outpatient management were provided the following day. The first NDC appointment was scheduled before discharge from the ED or hospital to guarantee rapid follow-up.

**Figure 1.**
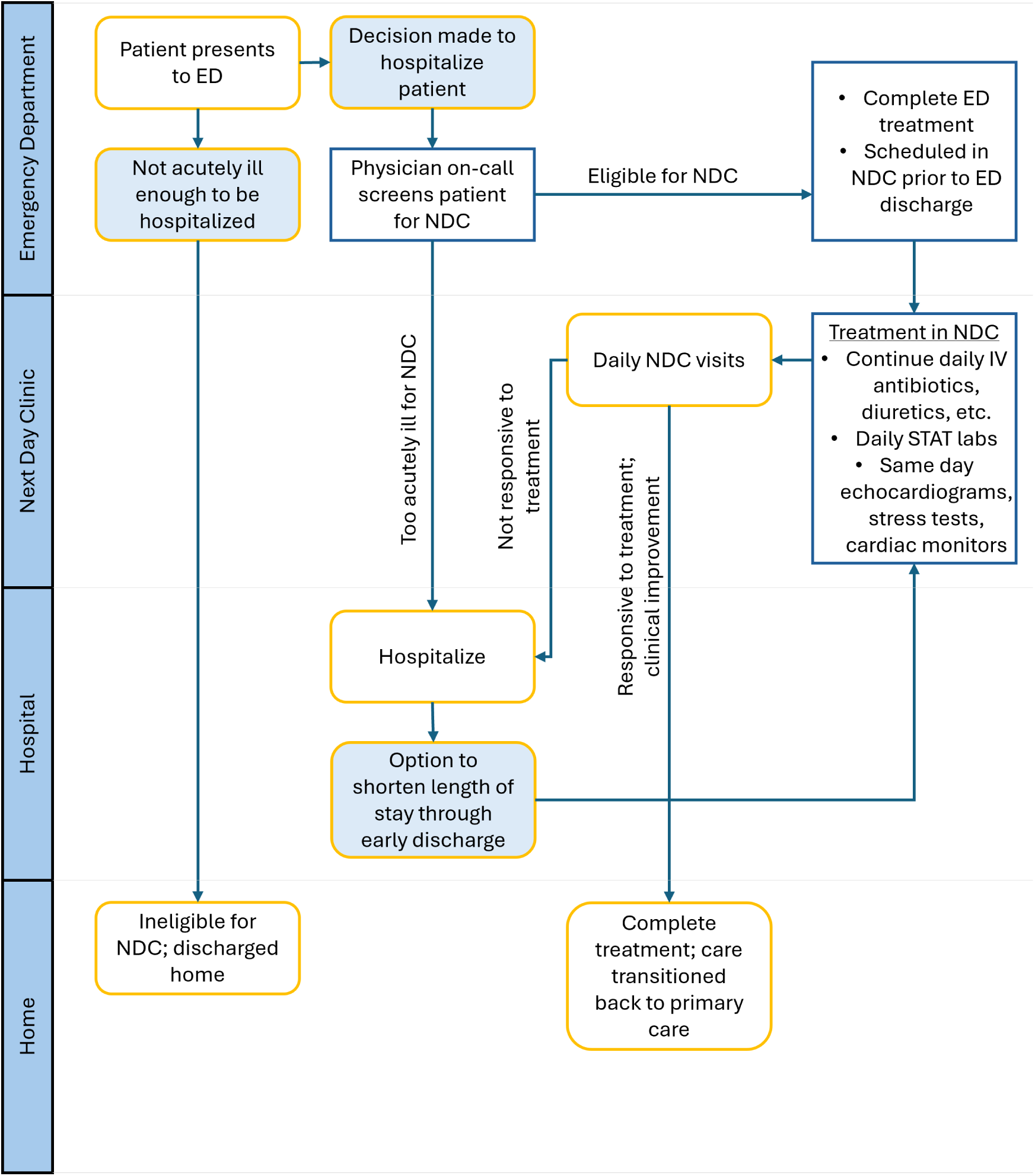
Olive View-UCLA Next Day Clinic (NDC) Referral and Treatment Workflow. Referral and treatment are divided into four swim lanes where the actions occur: emergency department (ED), Next Day Clinic (NDC), hospital, and home. Hospital admission avoidance referrals were those made in the ED, whereas early discharge referrals were those made in the hospital with the intent of shortening length of stay. Both types of referrals were denied if patients would have discharged home regardless of NDC follow-up. Eligible patients received hospital-level therapies in the outpatient NDC—including intravenous antibiotics or diuretics, same-day laboratory and imaging studies, and daily reassessment—until clinical resolution. Patients who clinically did not improve during NDC treatment were sent back to the ED or directly admitted to the hospital.

#### Inclusion and Exclusion Criteria

Each condition had a specific list of inclusion and exclusion criteria (Supplement 2), which were generally superseded by clinical judgment. The only general inclusion criterion was that patients were enrolled in or eligible for Department of Healthcare Services (DHCS) Medi-Cal (California’s Medicaid). General exclusion criteria across pathways included clinical instability (e.g., systolic blood pressure <90 mmHg), need for ICU-level monitoring or procedural/surgical intervention, poor social conditions deemed to be a barrier to reliable follow-up, severely immunocompromised, or pregnant.

### The NDC Model

The Olive View-UCLA NDC is a dedicated clinical service co-located with the hospital’s main Urgent Care Clinic, allowing it to launch with minimal startup costs by leveraging existing resources and infrastructure. One room of the Urgent Care Clinic is designated for NDC, and the clinic is staffed by a dedicated hospitalist or nurse practitioner with additional support from co-located urgent care physicians as needed. The NDC offers a variety of high-acuity services typically rendered in a hospital-based setting (Table 1), including but not limited to: daily IV antibiotics and diuretics, daily STAT labs, same-day echocardiograms and Ziopatch placement, same-day or next-day stress testing, same-day diagnostic radiology, same-day subspecialist consultation. Patients returned to the NDC every 24 hours to continue receiving services until they were out of their acute illness episode, as determined by the NDC provider.

**Table 1.**
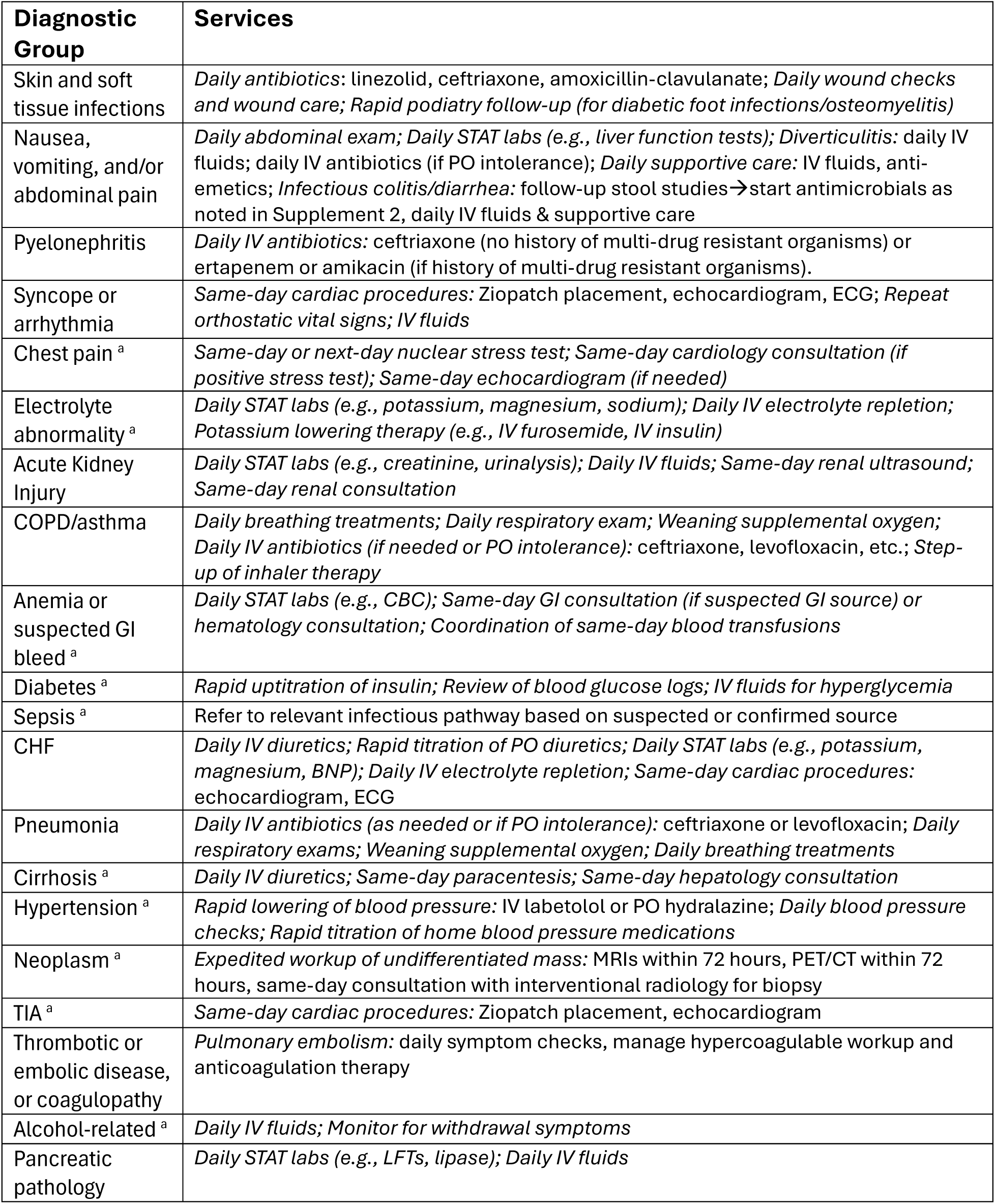
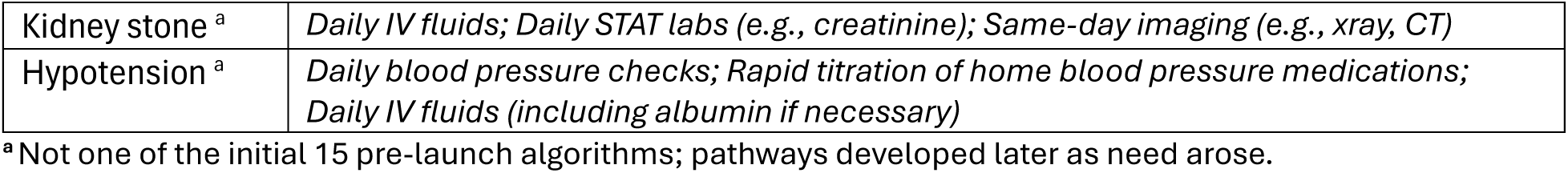
Abbreviated summary of next day clinic services by diagnostic group.

Prior to program launch, we designed 15 clinical treatment pathways utilizing these services in consultation with medical subspecialists: pyelonephritis, pneumonia, chronic obstructive pulmonary disease (COPD) or asthma exacerbation, congestive heart failure (CHF) exacerbation, syncope, cellulitis, diabetic foot ulcer or osteomyelitis, acute kidney injury (AKI), abdominal pain and vomiting, diverticulitis, infectious colitis, non-infectious colitis, pancreatitis, and pulmonary embolism (Supplement 2). After program launch, an additional 12 pathways were developed *ad hoc* based on need, for a total of 27 (eTable 2 in Supplement 1).

### Data and Outcomes

The primary outcome was days alive and out of hospital (DAOH)^18^ in the 30 days following the index ED visit. This was calculated as 30 days minus the number of days spent in a hospital under inpatient or observation status. Death during the 30-day period truncated the maximum DAOH at the death date minus the index ED visit date. Secondary outcomes were the number of hospital bed-days saved and the following events at 30-days from the index ED visit: all-cause ED revisits, all-cause unplanned hospitalization (after the index hospitalization for the matched control cohort), and all-cause mortality. Illness episodes were defined as the time from the index ED visit to the time of discharge from the hospital (control cohort) or the NDC (treatment cohort).

### Statistical Analysis

#### Matching

To construct a comparable control cohort, we matched patients referred to the NDC to patients seen in the ED in the 12 months prior to the program’s launch, with the goal of identifying patients who *could have* been referred to the NDC had it existed (Figure 2). The control pool was first limited to encounters for patients with eligible insurance, and with laboratory and vital sign values that fell within the observed range of the NDC cohort. The propensity score (the predicted probability of receiving an NDC referral) was then estimated using a logistic regression model.

**Figure 2.**
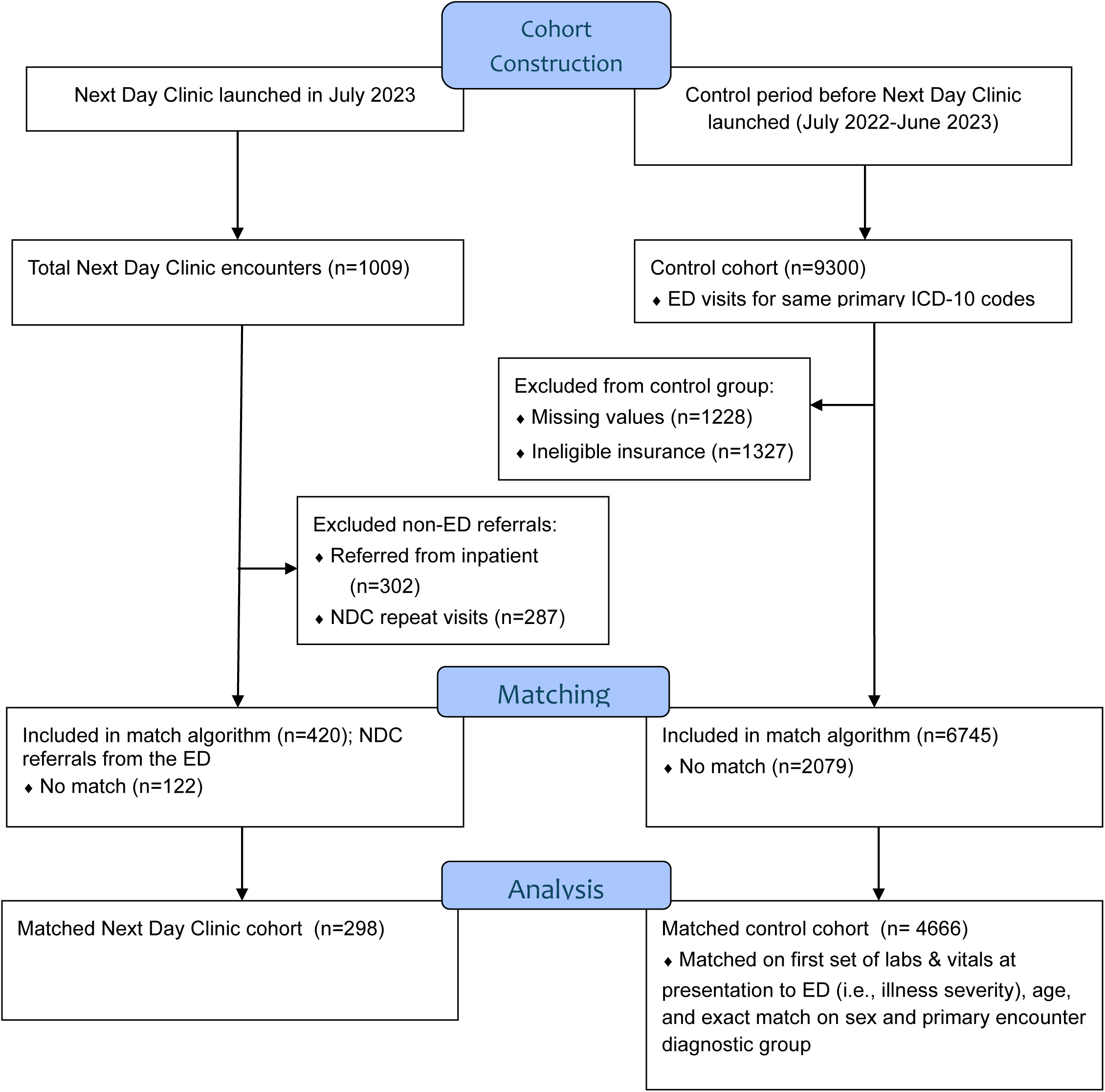
Cohort Flow Diagram. Of 1009 total NDC encounters during the 13-month study period, 420 (42%) were referrals from the ED. After exclusions and exact matching on age, sex, diagnostic group, and initial laboratory and vital-sign values, 298 ED-referred NDC patients were matched to 4666 ED encounters from the year prior to NDC launch. Matching to historical controls identified patients who would have been referred to the NDC, had it existed. This approach reduced bias arising from a contemporaneous control cohort, which by definition would have excluded suitable matches for NDC patients since eligible patients would have already been sent to the NDC. The flow diagram shows exclusions for missing data, ineligible insurance, and unmatched encounters, yielding the final analytic cohorts.

Matching was conducted on age, sex, the first complete set of labs and vitals obtained in the ED as a marker of presenting illness severity (all 14 labs and vitals are listed in eTable 1 in Supplement 1), and an exact match on the primary diagnosis group.^20^ We used the MatchIt package in R with optimal full matching,^19^ and encounters with missing variables were dropped (Figure 2).

#### Days alive and out of hospital (DAOH)

We performed the primary analysis on the matched cohorts using a weighted linear regression model to estimate the adjusted mean difference in DAOH. The model included an indicator for treatment group (NDC versus hospitalized), adjusted for all variables used in the matching, and applied weights derived from the optimal full matching. To account for multiple encounters from the same patient, we calculated cluster-robust standard errors at the patient level.^21,22^

#### Hospital bed-days saved

To estimate the number of bed-days saved from hospital admission avoidance referrals, we multiplied the mean difference in DAOH by the number of NDC referrals from the ED. Even though we aggressively screened patients to accept only those who would have been admitted in the absence of the NDC, we recognize the limitations in establishing a clear counterfactual of hospitalization in an observational study. To address this, we presented these estimates as a bounding exercise, with the most conservative estimate (lower bound) assuming only those patients meeting InterQual criteria^23^ were truly avoided hospitalizations. At OVMC, ED physicians intending to admit patients order the ED case manager to apply InterQual criteria to determine if the patient meets inpatient level of care, thereby establishing that patients intervened on and referred to NDC after InterQual orders would have been admitted in the absence of the NDC. The most liberal estimate (upper bound) assumed that 80% of ED referrals avoided hospitalizations regardless of InterQual orders (i.e., 20% of NDC referrals were for patients who would have been discharged anyways). This assumption was based on the observation that many ED providers recognized NDC candidates early in the ED encounter, and thus never placed an InterQual order even if they would have admitted patients in the absence of the NDC.

To further add to the conservative estimates of program impact, our analysis excluded early discharge referrals (i.e., only included hospital admission avoidance referrals from the ED) since we could not reliably determine if early discharge referrals truly shortened length of stay, as opposed to being used to expedite post-discharge follow-up for patients who were already discharging.

#### Sensitivity analyses

##### Alternative clustering

The linear regression to estimate mean DAOH difference was repeated using diagnostic group to define clusters rather than patient-level clustering.

##### Alternative modeling

Given that the DAOH outcome was right-skewed, quantile regression was used to estimate mean DAOH difference between groups. To account for the possibility that the effect of the NDC on DAOH might not be constant across encounters, we also implemented augmented inverse probability weighting (AIPW) within the double machine learning framework (using random forest).^24^

## RESULTS

During the 13-month study period, there were a total of 1009 NDC encounters. The mean age of NDC patients was 54 years (SD, 14.6 years) years, 44% were female, and the population was predominantly Latino/Hispanic (77%) with a reported primary language of Spanish (63%). Referral sources to the NDC were as follows: 420 (42%) from the ED, 302 (30%) from a hospitalist service, 287 (28%) were repeat NDC visits to continue treating the same illness.

By the end of the study period, the monthly show-rate was 88%. Compared to the year before NDC implementation, the annual mean cycle time per encounter of the co-located Urgent Care Clinic (as a marker of NDC impact on Urgent Care operations) increased by a non-statistically significant 2.4 minutes per encounter (p=0.68). Launching the NDC did not require any new clinical overhead, infrastructure, nursing, or administrative support since this was all shared with the Urgent Care Clinic. The only new resource required to launch the NDC was physician time at 0.25 FTE and nurse practitioner time at 0.75 FTE to staff 28 appointment slots per week, valued at $250,000.

### Propensity matching

To construct the control cohort, there were 9300 ED encounters for the eligible diagnosis groups from July 1, 2022-June 30, 2023. After exclusions for missing data and ineligible insurance, performing an exact match on diagnostic group, and additional matches on age, sex, and the presenting set of labs and vitals, we excluded 4634 (50%) of ED encounters (Figure 2).

From the 420 hospital admission avoidance NDC referrals, 122 (29%) did not have a suitable match with the control cohort so were excluded. This yielded an analytical cohort of 298 matched NDC patients and 4666 matched control patients from the year prior to NDC implementation.

The matched cohorts were statistically similar in terms of age, sex, race (Table 2) as well as presenting illness severity as determined by the first set of labs and vitals obtained in the ED (eTable 1 in Supplement 1). The case-mix of conditions is noted in eTable 2 in Supplement 1.

**Table 2.** Sample Characteristics.

| *Characteristic* | *Total NDC*<br>*Cohort (n=1009)* | *Matched NDC*<br>*cohort (n=298)* | *Matched*<br>*control cohort*<br>*(n=4666)* | *p value*<br>*(matched*<br>*cohorts)* |
| --- | --- | --- | --- | --- |
| ***Patient age, mean (SD)*** | 54.4 (14.6) | 54.52 (14.89) | 53.32 (15.22) | 0.52 |
| ***Patient gender, No. (%)*** |  |  |  |  |
| *Female* | 448 (44.4%) | 143 (48.0%) | 2019 (43.3) | 0.14 |
| *Male* | 561 (56.6%) | 155 (52.0%) | 2647 (56.7) | 0.30 |
| ***Patient race and ethnicity, No. (%)*** |  |  |  |  |
| *Asian or Pacific Islander* | 53 (5.3%) | 13 (4.4%) | 202 (4.3%) | 0.98 |
| *Black or African*<br>*American* | 37 (3.7%) | 13 (4.4%) | 184 (3.9%) | 0.72 |
| *Latino or Hispanic* | 774 (76.7%) | 232 (77.9%) | 3424 (73.4%) | 0.09 |
| *Non-Hispanic White* | 55 (5.5%) | 36 (12.1%) | 732 (15.7%) | 0.1 |
| *Other or multiple* | 90 (8.9%) | 4 (1.3%) | 124 (2.7%) | 0.17 |
| ***Payor type*** |  |  |  |  |
| *Medi-Cal* | 751 (74.4%) | 209 (70.1%) | 3508 (75.1%) | 0.31 |
| *Medi-Cal, Medicare*<br>*dual-eligible* | 121 (11.9%) | 36 (12.1%) | 751 (16.1%) | 0.1 |
| *Commercial* | 0 | 0 | 0 | n/a |
| *Uninsured* | 137 (13.6%) | 47 (15.8%) | 411 (8.8%) | <0.001 |
| ***NDC Statistics*** |  |  |  |  |
| *Referred from ED* | 420 (41.6%) | 298 (100%) | n/a | n/a |
| *Referred from inpatient* | 302 (30.0%) | 0 | n/a | n/a |
| *NDC return visit* | 287 (28.4%) | 0 | n/a | n/a |
| *NDC visits per index*<br>*encounter, mean (SD)* | 1.30 (0.66) | 1.43 (0.80) | n/a | n/a |

### Primary outcome

Unadjusted mean DAOH at 30-days from the ED visit was 29.5 days (SD, 2.3 days) in the NDC cohort, compared to 24.9 days (SD, 5.5 days) in the control cohort. The mean difference in DAOH adjusting for age, sex, primary diagnosis, and presenting illness severity was an additional 3.85 DAOH per encounter in the NDC group (SE 0.20 days, p<0.001). The sensitivity analyses produced similar estimates for mean DAOH difference, with the exception of the quantile regression which produced a more conservative estimate of +2.76 (0.14) DAOH in the NDC cohort (eTable 3 in Supplement 1).

### Secondary outcomes

Among the 298 matched NDC patients, 93 (31%) had InterQual applied to them in the ED and met inpatient criteria. To produce the most conservative estimate of hospital bed-days avoided (i.e., lower bound), we assumed that only these 93 NDC referrals avoided hospitalization, which at an average mean DAOH difference of 3.85 days (and no mortality) imputes a minimum of 358 hospital bed-days saved. Assuming that 80% of all NDC referrals from the ED avoided a hospitalization (i.e., 336), that yields an upper bound estimate of 1294 hospital bed-days saved. As noted previously, these estimates are conservative in that they do not account for bed-days avoided from shortened length of stay through the NDC early discharge pathway, which accounted for 30% of NDC referrals.

There were higher rates of 30-day ED revisits per 100 encounters in the NDC cohort compared to the control cohort (20.5 versus 13.0 per 100 encounters, p<0.001), but lower rates of both unplanned 30-day hospital readmission (5.7 versus 11.0 per 100 encounters, p<0.001) and all-cause 30-day mortality (0% versus 0.9%, p<0.001; Table 3).

**Table 3.** Cohort Outcomes.

|  | *Total NDC*<br>*Cohort*<br>*(n=1009)* | *Matched NDC*<br>*cohort (n=298)* | *Matched control*<br>*cohort (n=4666)* | *P-values*<br>*(comparing*<br>*matched groups)* |
| --- | --- | --- | --- | --- |
| ***Days alive and out of hospital (DAOH) at 30d from index ED visit*** |  |  |  |  |
| *DAOH, unadjusted, mean (SD)* | 29.4 (2.3) | 29.5 (2.3) | 25.2 (5.3) | p<0.001 |
| *DAOH, adjusted, difference versus matched control (SE)* | n/a | +3.85 (0.20) | n/a | p<0.001 |
| ***Unplanned utilization and events within 30d of index ED visit*** |  |  |  |  |
| *ED visits per 100 encounters* | 10.6 (0.97) | 20.5 (2.4) | 13.0 (0.49) | p<0.001 |
| *Hospital readmissions per 100 encounters* | 4.4 (0.64) | 5.7 (1.4) | 11.0 (0.46) | p<0.001 |
| *Mortality, all-cause (No., % of patients)* | 2 (0.2%) | 0 (0.0%) | 42 (0.9%) | p<0.001 |

## DISCUSSION

In this matched cohort study within U.S.’ second largest public safety-net health system, we found that a high-acuity Next Day Clinic was a feasible and safe alternative to traditional hospitalization for select, acutely ill adult patients referred from the ED. Treatment in the NDC was associated with an average of 3.85 more days alive and out of the hospital over a 30-day period per referral, compared to hospitalization as-usual. Over the course of the study, this amounted to as many as 1294 hospital bed-days saved. Importantly, this significantly lower in-hospital time was achieved without an associated increase in all-cause 30-day readmissions or mortality, suggesting the model is a safe substitute for hospital-based care in this population.

These findings position the NDC as a distinct and valuable innovation in the evolving landscape of acute care delivery. The model’s primary contribution is in its creation of a crucial intermediate level of care between hospitalization and traditional outpatient acute care using primary care or walk-in clinics. The NDC provides ED physicians with a middle ground between these two dispositions, serving as a practical alternative to hospitalization by offering services historically rendered in a hospital setting: multi-day courses of IV antibiotics and diuretics, daily STAT laboratory monitoring, and rapid access to advanced imaging and cardiodiagnostics, as examples. Because the NDC pathways are designed for patients who often have an expected length of stay of only a few days, many of whom might otherwise be admitted under observation status, the NDC effectively functions as a first-of-its-kind “Ambulatory Observation Unit.” This stands in contrast to traditional ED or hospital-based observation units by recreating many of the services that these offer but in a pre-planned, potentially multi-day course of treatment in an ambulatory setting.^7,25,26^

The NDC model is also differentiated from other hospital avoidance strategies by centralizing care within existing infrastructure, which maximizes economies of scale and clinical comprehensiveness. This approach contrasts with other important models, including Hospital at Home, remote patient monitoring without Hospital at Home, and specialized outpatient programs. While Hospital at Home is effective at delivering select inpatient-level services,^9,27^ it faces substantial barriers to scalability and sustainability. The model is resource-intensive, with nurses often limited to seeing 4-6 patients per day,^15,16^ and its high startup costs of $1.5-2 million have contributed to its adoption by fewer than 300 U.S. hospitals.^5,12–14^ Furthermore, its reliance on a federal reimbursement waiver, which lapsed as of September 30, 2025 causing numerous Hospital at Home programs to fail, raises concerns about its long-term viability.^17^ In contrast, the NDC model leverages existing resources by co-locating with an urgent care, allowing it to be launched with only 1.0 FTE (valued at under $300,000), and maximizing economies of scale since there is no travel time between patients and providers have full access to STAT diagnostics and imaging. Importantly, the NDC providers bill as a standard urgent care, making the model financially viable without relying on uncertain reimbursement waivers. While many remote patient monitoring programs are effective for post-discharge monitoring,^28^ they are not designed to deliver high-acuity interventions such as daily IV therapies or rapid advanced diagnostics, since they rely on *ad hoc* clinic visits and a majority of patients do not utilize these.^10^ Finally, specialized programs like Outpatient Parenteral Antimicrobial Therapy (OPAT) or chest pain pathways centralize care but are limited by a narrow clinical focus, restricting their scalability to the diverse range of conditions leading to hospitalization.^29,30^ The NDC fills an important niche among these other valuable programs by offering a highly economical, scalable, and clinically robust alternative for the substantial portion of acutely ill patients who require many of the services rendered in a hospital, and are too sick to rely on *ad hoc* or episodic clinic visits.

The operational and policy implications of this model are substantial. For hospital administrators facing severe capacity constraints, the NDC allows for both ED and hospital unit decompression, which is especially important given the U.S.’ looming national hospital bed shortage.^1^ For payors, each avoided hospitalization through the NDC replaces a costly inpatient stay with clinic visits that are several orders of magnitude less expensive, savings that could theoretically be passed on to patients in the form of lower premiums. From the patient perspective, the NDC allows them to stay home with loved ones, avoiding the risks of psychological distress, financial toxicity, and hospital-associated adverse events.^31,32^

While a formal cost effectiveness analysis was not within the scope of this study, the potential cost savings of the NDC model are enormous. The actual mean cost per day of hospitalization at a Los Angeles County DHS hospital in 1997-1998 was $1,483, which inflation adjusted is $2,954 per day of hospitalization in 2025 dollars.^33^ Conservatively estimating that each hospital bed-day costs $2,500 to the organization, the estimate of 358-1294 hospital bed-days saved imputes $895,000-$3.24M in avoided hospital expenditures over the 13 month period.

### Limitations

The primary limitation of this study is that its retrospective observational design is susceptible to selection bias. Although we matched on a comprehensive set of 17 clinical variables that included the first set of labs and vitals in the ED (as a measure of presenting illness severity), we cannot account for unmeasured confounding from factors not discretely captured in the EHR such as social support, health literacy, or motivation. By reporting the estimated effect of the NDC as an upper and lower bound, we minimize the possibility of an inflated effect size since the lower bound is calculated from NDC patients who have a clear counterfactual of hospital admission were it not for the NDC intervention (i.e., they had an InterQual order prior to NDC referral). This lower bound represents the minimum effect of the NDC, since it mitigates the possibility that a referred patient would have been discharged from the ED even without the NDC. We took numerous other measures to avoid overestimating the effect of the NDC, such as excluding from our analysis early discharge referrals from the inpatient wards, since there was no way to establish a counterfactual that they would have remained hospitalized in the absence of the NDC.

Second, we cannot exclude the possibility that patients received care outside of the Los Angeles County DHS system during the follow-up period which may have over- or under-estimated the outcomes in one or both cohorts. However, this is unlikely given that the study population was restricted to those exclusively with DHS insurance (i.e., Medi-Cal) among whom there is very little leakage outside the safety net.^34^ Third, this was a single center evaluation, so further studies are needed to establish generalizability of the NDC. Finally, this analysis was not designed to collect patient-reported outcomes such as satisfaction with care, quality of life, and functional status, which will be needed for a comprehensive programmatic evaluation.

## Conclusion

The Next Day Clinic model is a feasible and promising strategy for replacing hospitalization among select acutely ill patients being admitted through a safety-net ED. By providing pre-planned, potentially multi-day courses of IV therapies and access to rapid STAT labs and advanced imaging/cardiodiagnostics, this model was associated with 358-1294 hospital bed-days saved, which translates to $895,000-$3.24M in avoided hospital healthcare expenditures. The NDC may be an important new care model for mitigating rising hospital occupancies and healthcare spending and warrants further prospective evaluations.

## Data Availability

All data produced in the present study are available upon reasonable request to the authors pending a data use agreement with the Los Angeles County-Department of Health Services.

