## Supplement 1 for "Evaluation of a High-Acuity Next Day Clinic for Hospital Admission Avoidance: A 13-Month Cohort Study"

### Supplementary Appendix 1 for:

Evaluation of a High-Acuity Next Day Clinic for Hospital Admission Avoidance: A 13-Month Cohort Study

**eTable 1**. Laboratory and vital sign values at index ED presentation for the propensity matched cohorts, as a marker of presenting illness severity.

| *Value* | *Matched NDC Cohort (n=298)* | *Matched control cohort (n=4666)* | *p-value* | *Included in matching algorithm?* |
|---|---|---|---|---|
| ***SpO2 (mean (SD))*** | 98.05 (2.05) | 97.63 (2.62) | 0.01 | Yes |
| ***RespRate (mean (SD))*** | 18.07 (1.83) | 18.73 (2.23) | <0.001 | Yes |
| ***BP_Sys (mean (SD))*** | 137.06 (24.62) | 134.87 (23.58) | 0.12 | Yes |
| ***BP_Dia (mean (SD))*** | 76.52 (15.97) | 78.59 (16.28) | 0.03 | Yes |
| ***WBC (mean (SD))*** | 8.61 (4.80) | 9.82 (4.84) | <0.001 | Yes |
| ***Hgb (mean (SD))*** | 12.25 (2.49) | 12.41 (2.43) | 0.28 | Yes |
| ***Platelet (mean (SD))*** | 265.34 (110.21) | 268.47 (112.97) | 0.64 | Yes |
| ***Sodium (mean (SD))*** | 135.44 (3.67) | 135.04 (3.68) | 0.07 | Yes |
| ***Potassium (mean (SD))*** | 4.13 (0.59) | 4.17 (0.55) | 0.24 | Yes |
| ***Chloride (mean (SD))*** | 101.92 (4.93) | 100.64 (4.82) | <0.001 | Yes |
| ***CO2 (mean (SD))*** | 24.15 (3.16) | 24.25 (3.13) | 0.58 | Yes |
| ***BUN (mean (SD))*** | 19.13 (14.01) | 17.92 (13.29) | 0.13 | Yes |
| ***Creatinine (mean (SD))*** | 1.25 (1.04) | 1.40 (1.60) | 0.11 | Yes |
| ***Glucose (mean (SD))*** | 150.69 (98.54) | 153.08 (91.98) | 0.66 | Yes |
| ***Sed.Rate (mean (SD))*** | 35.36 (27.45) | 36.23 (27.40) | 0.91 | No |
| ***INR (mean (SD))*** | 1.21 (0.36) | 1.18 (0.31) | 0.30 | No |
| ***Albumin (mean (SD))*** | 3.48 (0.69) | 3.48 (0.67) | 0.92 | No |
| ***ALT (mean (SD))*** | 58.83 (276.50) | 44.87 (77.60) | 0.08 | No |
| ***AST (mean (SD))*** | 55.51 (186.70) | 48.91 (94.62) | 0.42 | No |
| ***Bilirubin (mean (SD))*** | 1.61 (3.84) | 1.33 (2.11) | 0.12 | No |
| ***hs.CRP (mean (SD))*** | 56.23 (75.20) | 71.95 (79.05) | 0.30 | No |
| ***Lipase.Lvl (mean (SD))*** | 52.23 (66.48) | 45.09 (52.90) | 0.23 | No |
| ***Lactate.Lvl (mean (SD))*** | 1.54 (0.78) | 1.66 (0.74) | 0.34 | No |
| ***Magnesium (mean (SD))*** | 2.03 (0.27) | 2.01 (0.27) | 0.64 | No |
| ***Troponin.I (mean (SD))*** | 0.01 (0.02) | 0.02 (0.03) | 0.02 | No |
| ***BNP (mean (SD))*** | 396.62 (756.87) | 553.86 (870.13) | 0.11 | No |

**eTable 2**. Diagnostic group case-mix of the cohorts.

| | *NDC matched ED Referrals (n=298)* | | *NDC all ED Referrals (n=420)* | | *NDC all referrals (n=1009)* | | *Matched ED cohort (n=4666)* | |
|---|---|---|---|---|---|---|---|---|
| ***Condition*** | Count | % | Count | % | Count | % | Count | % |
| *SSTI* | 36 | 12.1 | 46 | 11.0 | 68 | 6.7 | 471 | 10.1 |
| *Nausea, vomiting, and/or abdominal pain* | 33 | 11.1 | 47 | 11.2 | 94 | 9.3 | 569 | 12.2 |
| *Pyelonephritis* | 33 | 11.1 | 46 | 11.0 | 79 | 7.9 | 464 | 9.9 |
| *Syncope* | 32 | 10.7 | 45 | 10.7 | 74 | 7.4 | 330 | 7.1 |
| *Chest pain* | 23 | 7.7 | 33 | 7.9 | 54 | 5.4 | 377 | 8.1 |
| *Electrolyte* | 19 | 6.4 | 27 | 6.4 | 32 | 3.2 | 270 | 5.8 |
| *AKI* | 19 | 6.4 | 25 | 6.0 | 43 | 4.3 | 321 | 6.9 |
| *COPD/asthma* | 18 | 6.0 | 26 | 6.2 | 75 | 7.5 | 317 | 6.8 |
| *Anemia* | 15 | 5.0 | 20 | 4.8 | 40 | 4.0 | 181 | 3.9 |
| *Diabetes* | 14 | 4.7 | 21 | 5.0 | 42 | 4.2 | 247 | 5.3 |
| *Sepsis* | 11 | 3.7 | 15 | 3.6 | 38 | 3.8 | 191 | 4.1 |
| *CHF* | 9 | 3.0 | 11 | 2.6 | 19 | 1.9 | 149 | 3.2 |
| *Pneumonia* | 8 | 2.7 | 12 | 2.9 | 39 | 3.9 | 177 | 3.8 |
| *Cirrhosis* | 5 | 1.7 | 7 | 1.7 | 28 | 2.8 | 135 | 2.9 |
| *HTN* | 5 | 1.7 | 5 | 1.2 | 21 | 2.1 | 60 | 1.3 |
| *GI bleed* | 5 | 1.7 | 3 | 0.7 | 76 | 7.6 | 130 | 2.8 |
| *Arrythmia* | 5 | 1.7 | 8 | 1.9 | 18 | 1.8 | 74 | 1.6 |
| *Neoplasm* | 4 | 1.3 | 4 | 1.0 | 10 | 1.0 | 97 | 2.1 |
| *TIA* | 2 | 0.7 | 0 | 0.0 | 8 | 0.8 | 0 | 0.0 |
| *Thrombotic or embolic disease* | 1 | 0.3 | 3 | 0.7 | 7 | 0.7 | 55 | 1.2 |
| *Alcohol* | 1 | 0.3 | 3 | 0.7 | 6 | 0.6 | 51 | 1.1 |
| *Not specified* | 0 | 0.0 | 13 | 3.1 | 62 | 6.2 | 0 | 0.0 |
| *Pancreatic pathology* | 0 | 0.0 | 0 | 0.0 | 4 | 0.4 | 0 | 0.0 |
| *Kidney stone* | 0 | 0.0 | 0 | 0.0 | 6 | 0.6 | 0 | 0.0 |
| *Diverticulitis* | 0 | 0.0 | 0 | 0.0 | 32 | 3.2 | 0 | 0.0 |
| *Coagulopathy* | 0 | 0.0 | 0 | 0.0 | 21 | 2.1 | 0 | 0.0 |
| *Hypotension* | 0 | 0.0 | 0 | 0.0 | 13 | 1.3 | 0 | 0.0 |
| | 298 | 100.0 | 420 | 100 | 1009 | 100.0 | 4666 | 100.0 |

**eTable 3.** Results of regressions included in sensitivity analyses.

| ***Regressions*** | ***Difference in days alive and out of hospital (DAOH) between the Next Day Clinic and matched control cohort*** | ***p-value*** |
|---|---|---|
| ***Linear, heteroskedasticity-consistent SEs (primary analysis)*** | +3.85 (0.20) | p<0.001 |
| ***Linear, cluster robust SEs (patient)*** | +3.85 (0.20) | p<0.001 |
| ***Linear, cluster robust SEs (diagnostic group)*** | +3.85 (0.20) | p<0.001 |
| ***Quantile, cluster bootstrap at patient level*** | +2.76 (0.14) | p<0.001 |
| ***Double machine learning*** | +3.86 (0.28) | p<0.001 |

*Abbreviations:* SE, standard error.
