## Supplement 2 for "Evaluation of a High-Acuity Next Day Clinic for Hospital Admission Avoidance: A 13-Month Cohort Study"

### Supplementary Appendix 2 for:

Evaluation of a High-Acuity Next Day Clinic for Hospital Admission Avoidance: A 13-Month Cohort Study

Clinical Treatment Protocols

### CELLULITIS (excluding diabetic foot infections)

**Inclusion criteria for next day clinic:**

- Any skin/soft tissue infection that does not meet the exclusion criteria
- Cellulitis that "failed" oral cephalosporins as outpatient are great candidates

**Illness-specific exclusion criteria for next day clinic (in addition to general exclusion criteria):**

- Diabetic foot infection-- refer to that algorithm instead
- Necrotizing skin/soft tissue infection
- Evidence of sepsis by qSOFA criteria (RR≥22 breaths/min, altered mental status (GCS<15), SBP<100 mmHg)
- Treatment with IV antibiotics of the same site within the prior month
- Cellulitis of the face, hands, or overlying joints
- History of limb amputation or complex skin/soft tissue infections in the past
- Severe penicillin allergy (e.g., SJS/TENS), non-severe PCN allergy is okay

**Treatment in ED**

1. Use sharpie to outline borders of affected area, and follow the appropriate pathway below
2. If cellulitis is the result of an animal/human bite, skip to that section. Otherwise proceed to *Step 3*
3. If purulent infection, ensure I&D is performed in ED and sent for culture (skip if non-purulent)
4. After giving appropriate IV antibiotics in the ED, set up the following dispo antibiotics:
    - For standard patients: linezolid 600mg PO BID x7 days
    - For patients with contraindication to linezolid: administer 1.2g oritavancin IV x1
        - Contraindications include: h/o poor compliance or follow-up, concurrent use of pseudoephedrine, dopaminergic agents, SSRIs, TCAs, triptans, buspirone
        - Can be given without ID approval: contact on-call pharmacist
5. Proceed to *Schedule Follow-up*

*For Animal/Human Bite*

1. Assuming patient has received ampicillin-sulbactam, schedule another 3g dose 4-6 hours after last
    - Discharge with amoxicillin-clavulanate 875mg q8h x7 days
2. Proceed to *Schedule Follow-up*

*Schedule Follow-up*

1. Schedule NDC appointment within 48-72 hours for wound/symptom check and to follow-up on cultures (if any sent).
    a. Bites can be brought back within 24 hours for additional doses of Unasyn

*Dosing must be adjusted for renal impairment unless otherwise specified

**Next day clinic workflow:**

*First follow-up (48-72 hours after ED visit), in-person or telehealth*

1. Assess for any of the following red flags: Any hospitalizations at OV or any other hospital, or any unscheduled urgent care or ED visits anywhere, for any cause (record visit dates and length of hospitalization if any). Any new/worsening fever, chills, malaise, nausea, vomiting, tenderness/pain at affected area, or enlargement of affected area (compared to sharpie outline).
    a. If worsening symptoms, consider abscess and appropriate imaging (US or CT)
2. Review culture results from ED if any were sent, and adjust antibiotics if appropriate
3. *Bites only (otherwise skip to Step 4): Administer 3g ampicillin-sulbactam IV x1*
4. Determine disposition/follow-up:
    a. If improving well no follow-up needed (PCP can assume care)
    b. If marginal improvement can schedule for NDC follow-up based on clinical judgment

*All additional follow-ups will follow the same steps as above*

### SYNCOPE

**Exclusion criteria for next day clinic:**

- Syncope definitively due to or high clinical suspicion for stroke, TIA, or seizure
- Family history of sudden cardiac death
- Syncope due to symptomatic anemia or malignant arrhythmia
- Presence of typical angina
- Loss of consciousness >5 min
- Not at baseline mental status at time of evaluation
- Major trauma
- Intoxication with alcohol or other drugs

**Treatment in ED:**

1. Patient must be placed on continuous cardiac monitor if not already done
2. Ensure that troponin and ECG are completed. Head imaging only required if history and/or exam concerning for intracranial pathology. CT pulmonary angiogram is only required if there is suspicion for PE.
3. Review exclusion criteria, and if present stop algorithm and deliver care as usual.
4. Administer IV fluids using clinical judgment
5. If possible, obtain TTE in ED. Otherwise can obtain TTE in the Next Day Clinic
6. Calculate Canadian Syncope Risk Score (CSRS)-- calculate below, or scan QR Code:
    a. Vasovagal predisposition (warm/crowded place, prolonged standing, intense emotion/pain, etc.) (-1)
    b. History of heart disease (CAD, afib/flutter, CHF, valvular disease) (+2)
    c. Triage SBP<90 or >180 mmHg (+2)
    d. Elevated troponin >99th percentile for sex (+2)
    e. Abnormal QRS axis (< -30° or > 100°) (+1)
    f. QTc >480 ms (+2)
    g. Leading diagnosis vasovagal syncope (-2)
    h. Leading diagnosis cardiac syncope (+2)
7. Interpret CSRS as follows:
    a. Low = -3 to 0  2h of ED monitoring for arrhythmia and schedule 24-36 hour NDC follow-up, **in-person**
    b. Medium = 1 to 3  6h of ED monitoring and schedule 24-36 hour NDC follow-up, **in-person**
    c. If CSRS 4 to 11, then consider admission
    d. <u>Clinical judgment supersedes CSRS</u>

*Dosing must be adjusted for renal impairment unless otherwise specified

**Treatment in NDC:**

*First follow-up (~24 hours after ED visit), in-person*

1. Assess for any of the following red flags: Any hospitalizations at OV or any other hospital, or any <u>unscheduled</u> urgent care or ED visits anywhere, <u>for any cause</u> (record visit dates and length of hospitalization if any). Any syncope, presyncope, or light headedness/dizziness, palpitations.
2. Perform orthostatic vital signs
    -  If negative, proceed to step 3
    -  If positive, administer IV fluids based on clinical judgment and repeat
        -  If orthostatics remain positive, consider fludricortisone 0.05mg qday, and counsel patient on obtaining compression stockings
3. If ECG in the ED was abnormal  repeat ECG. Otherwise move to Step 4
    a. If arrhythmia is suspected cause of syncope, discuss disposition with inpatient cardiology consult
4. Auscultate carotids  If bruit then order carotid duplex. Otherwise move to Step 5.
5. If there is concern for structural heart disease (e.g., known structural pathology, physical exam), then obtain TTE if this was not performed in ED
6. If there is concern for arrhythmia as etiology of syncope, call cardiac device clinic to have same-day Ziopatch placed.
7. If there is concern for structural heart disease, call echo clinic to have same-day TTE completed.
8. No further NDC follow-up required (PCP can assume care)

*Dosing must be adjusted for renal impairment unless otherwise specified

### PYELONEPHRITIS

**Inclusion criteria for next day clinic:**

- Patients with UTIs (including pyelonephritis) <u>who would normally be admitted</u>
- History of MDR organisms (including ESBL) are acceptable
- Frequently recurrent UTIs are acceptable

**Illnes**s-specific exclusion criteria for next day clinic (in addition to general exclusion criteria):

- Patients with UTIs who would normally be directly discharged from the ED
- New onset oliguria or complete anuria
- Patient is bacteremic or unstable after initial treatment in ED
- Serum creatinine more than 3.0 times baseline
- Evidence of stones that are a suspected nidus of infection, or new onset hydronephrosis

Treatment in ED

1. Administer IV antibiotics in the ED as follows:
    a. History of, risk factors for, or known multi-drug resistant organisms (MDRO) in urine
        i. If Cr<1.5: Administer amikacin 15 mg/kg IV*
        ii. If Cr>1.5: Ertapenem 1g IV
    b. Low concern for MDRO
        i. Administer antibiotics as usual (e.g., ceftriaxone 1g IV)
2. If foley catheter or suprapubic tube is present, replace catheter and proceed down one of following algos

*Acute cystitis that would normally be admitted in the absence of the NDC*

1. Schedule patients in NDC within 24 hours to continue receiving IV antibiotics
2. Remember that any patient who doesn't require admission and would likely improve on oral antibiotics alone does not require NDC follow-up.

*Pyelonephritis that would normally be admitted in the absence of the NDC*

1. Schedule patients in NDC within 24 hours to continue receiving IV antibiotics
2. Remember that any patient who doesn't require admission and would likely improve on oral antibiotics alone does not require NDC follow-up.

*Dosing must be adjusted for renal impairment unless otherwise specified

OV-UCLA Next Day Clinic Inclusion Criteria and Treatment Algorithms
Updated: September 5, 2023

**Next day clinic workflow:**

*All follow-up visits follow this pathway:*

1. Assess for any of the following red flags: Any hospitalizations at OV or any other hospital, or any unscheduled urgent care or ED visits anywhere, for any cause (record visit dates and length of hospitalization if any). Any new/worsening urinary retention, hematuria, oliguria, anuria, flank pain, chills/rigors, fevers, sweats.
2. Administer IV antibiotics as follows:
    - If C&S data from ED has resulted, use those to guide therapy
    - If no C&S results yet, administer another dose of antibiotics as follows:
        - Amikacin 15 mg/kg IV* to cover all GNs (including ESBL + CRE), OR
        - If Cr >1.5 and concern for ESBL (any of the below), then give ertapenem 1g IV*, OR
        - If Cr >1.5 and no concern for ESBL, then give ceftriaxone 1g IV
3. Schedule follow-up:
    a. If no C&S results yet, schedule for in-person NDC within 24 hours for another dose of IV abx
    b. If C&S has resulted and patient on appropriate oral abx, no further NDC visits needed (PCP assumes care)

*Dosing must be adjusted for renal impairment unless otherwise specified

### DIABETIC FOOT ULCER AND/OR OSTEOMYELITIS

**Inclusion criteria for next day clinic:**

- Diabetic foot ulcer with or without concern for osteomyelitis
- Lower extremity osteomyelitis with or without overlying skin infection
- Any cellulitis in a patient with diabetes that does not meet the exclusion criteria

**Illness-specific exclusion criteria for next day clinic (in addition to general exclusion criteria):**

- Concern that patient will require surgical intervention beyond simple bedside debridement
- Necrotizing skin/soft tissue infection or wet gangrene
- Patient has unstable vitals and/or high suspicion for bacteremia
- Treatment with IV antibiotics of the same site within the prior month
- Severe penicillin allergy (e.g., SJS/TENS); non-severe PCN allergy is okay

**Treatment in ED**

1. Use sharpie to outline borders of affected area
2. Obtain ESR and CRP
3. Use clinical judgment when ordering xray/CT-- they are not required as part of the algorithm
4. If there is concern for necrotizing infection, wet gangrene, or acute need for surgery, stop algorithm and admit
5. If there is concern for overlying skin/soft tissue infection, proceed below. Otherwise skip to step 6.
    a. Give Ampicillin/sulbactam 3g* IV x1 regardless of if there is purulence
        - Discharge with amox/clav 875mg PO q12h* x7d (can get more IV abx in NDC)
        - If h/o MRSA, also give minocycline 200mg PO x1 (loading in ED), followed by 100mg PO q12h x7d
6. If no overlying skin/soft tissue infection, do not administer antibiotics in order to maximize culture yield
7. Proceed down one of the two options:

*1) If wound probes to bone OR xray/CT suspicious for osteomyelitis OR evidence of surrounding necrotic tissue:*

1. Consult to podiatry for debridement/deep wound culture/bone culture in ED
2. If podiatry consult unavailable in ED (e.g., after 5pm):
    a. Please send whatever deep wound culture can be obtained in ED
    b. Input patient information into Podiatry Consult Pool on Teams for urgent follow-up
    c. Per Dr. Nouvong, in addition to "B" instruct patient to report to podiatry clinic at next available time:
        - Monday or Tuesday at 7:30am
        - Thursday or Friday at 12:30pm

*2) If wound does not probe to bone AND xray/CT does not suggest osteomyelitis (if obtained) AND no necrotic tissue:*

1. Obtain deep wound tissue culture (not a simple swab), can be seen by podiatry within 1 week
2. Input patient information into Podiatry Consult Pool on Teams for non-urgent follow-up (within 1 week)

*Schedule Follow-up for all patients as follows:*

1. All patients should have podiatry follow-up as noted above.
2. If overlying cellulitis: Schedule Next Day Clinic in-person appointment within 24 hours for further Unasyn
3. If no overlying cellulitis: Schedule Next Day Clinic in-person within 2-5 days

*Dosing must be adjusted for renal impairment unless otherwise specified

**Treatment in NDC:**

*First follow-up (1-5 days after ED visit), in-person*

1. Assess for any of the following red flags: Any hospitalizations at OV or any other hospital, or any unscheduled urgent care or ED visits anywhere, for any cause (record visit dates and length of hospitalization if any). Any new/worsening fever, chills, malaise, nausea, vomiting, tenderness/pain at affected area, or enlargement of affected area (compared to sharpie outline).
2. If overlying cellulitis, give antibiotics as below. If no cellulitis then skip to *Step 3*.
    a. Use culture data to guide antibiotics if available
    b. If no culture data yet, given ampicillin-sulbactam 3g IV x1
3. Ensure that appropriate podiatry follow-up has been completed or is scheduled
    a. Per Dr. Nouvong, if podiatry f/u has happened, please ensure that these cultures are followed (either by NDC, podiatry, or PCP) and patient is started on appropriate abx
4. Determine follow-up
    a. If overlying cellulitis, can bring back as needed for further doses of ampicillin-sulbactam
    b. Patient can continue to follow in NDC until started on antibiotics or be transitioned back to PCP as appropriate.

OV-UCLA Next Day Clinic Inclusion Criteria and Treatment Algorithms
Updated: September 5, 2023

**DIVERTICULITIS**

**Inclusion criteria for next day clinic:**
- Acute uncomplicated diverticulitis
- CT scan indicating Hinchey I (peri-colonic/mesenteric thickening without fluid collections)
- Still able to drink ~4 glasses of water per day, and able to tolerate pills

**Illness-specific exclusion criteria for next day clinic (in addition to general exclusion criteria):**
- CT A/P with any “red flags” that require surgical evaluation (e.g., SBO) or are at high risk of decompensation (e.g., toxic megacolon).
- Complete or near-complete PO intolerance of liquids or pills
- CT scan indicating anything other than Hinchey I (any fluid collections, bubbles, purulence, etc.)
- WBC > 16,000
- Evidence of sepsis by qSOFA (2 out of 3: AMS, SBP≤100, RR≥22)
- QTc ≥ 500 ms
- Failure of PO antibiotics for current diverticulitis episode
- Diverticulitis episode within the last 6 months

**Treatment in ED:**
1. Order CRP if not already drawn
2. Review or obtain ECG to ensure QTc<500, administer IV ondansetron and/or metoclopramide as usual
   ▯ Can use 1-2mg lorazepam PO for nausea if refractory to ondansetron/metoclopramide
3. Ensure patient has been adequately rehydrated (e.g., 1-3L crystalloids), based on clinical judgment
4. If anaerobic coverage has not been given, administer metronidazole 500mg PO* x1
   a. Additionally give PO ciprofloxacin 500mg if not already administered
5. Standardized guidelines/suggestions for treating <u>abdominal pain</u>
   * In general, <u>use functional pain rather than reported pain (0-10 scale)</u> as more objective measure: if patient can use phone or sleep in ED then they do not need to be admitted. Should be gravely disabled from sx to admit.
   ▯ Give all patients PO tylenol 1000mg and IV methocarbamol 500-1000mg
   ▯ If no contraindications and low s/f PUD/gastritis, give ketorolac 30mg IM x1
   ▯ If opioids are absolutely needed, <u>consider oxycodone 5mg PO</u> (must tolerate PO to qualify for NDC)
6. If sx responding to above treatment (e.g., able to use phone/sleep; not gravely disabled) ▯ *Schedule Follow-up*
   ▯ If gravely disabled from pain, please consider another round of treatment and reassess in 1 hr. If still gravely disabled (can’t complete ADLs at home), then patient doesn’t qualify for NDC-- can admit patient.

*Schedule Follow-up*
1. Confirm that telephone and email (if they have one) are correct
2. Standardized guidelines/suggestions for treating <u>nausea/vomiting</u> at discharge:
   - Orally disintegrating ondansetron 8mg SL q8h prn x30d, +/- lorazepam 1mg PO q6h prn x5d
3. Standardized guidelines/suggestions for treating <u>abdominal pain</u> at discharge:
   - Consider PO methocarbamol, and alternating tylenol and ibuprofen (if no contraindications)
4. Option 1: If clinical judgment suggests additional IV fluids and/or medications (non-opioids) are indicated:
   ▯ Schedule Next Day Clinic **in-person** appointment within 24 hours
5. Option 2: If no further parenteral treatment or lab rechecks are necessary:
   ▯ Schedule Next Day Clinic **video or phone** appointment in 24-48 hours based on sx severity

*Dosing must be adjusted for renal impairment unless otherwise specified

**Next day clinic workflow:**

*First follow-up visit (24-48 hours), telehealth or in-person*

1. Assess for the following red flags: Any hospitalizations at OV or any other hospital, or any unscheduled urgent care or ED visits anywhere, for any cause (record visit dates and length of hospitalization if any). Intractable nausea/vomiting, worsening abdominal pain, fevers/chills/rigors, new/worsening hematochezia.
2. If in-person visit, can administer further IV fluids and PO ciprofloxacin 500 mg and PO metronidazole 500mg*
3. Schedule follow-up:
    a. If patient recovering rapidly without complications, no further NDC f/u required (PCP assumes care)
    b. If patient requires closer follow-up schedule in ~1-3 days by **video/phone** or **in-person**

*Second follow-up visit (OPTIONAL; 2-5 days after initial ED visit), telehealth or in-person*

1. Assess for the following red flags: Any hospitalizations at OV or any other hospital, or any unscheduled urgent care or ED visits anywhere, for any cause (record visit dates and length of hospitalization if any). Intractable nausea/vomiting, worsening abdominal pain, fevers/chills/rigors, new/worsening hematochezia.
2. If in-person visit, can administer further IV fluids and PO ciprofloxacin 500 mg and PO metronidazole 500mg*
3. No further NDC follow-up required (PCP can assume care)

*Dosing must be adjusted for renal impairment unless otherwise specified

OV-UCLA Next Day Clinic Inclusion Criteria and Treatment Algorithms
Updated: September 5, 2023

**ABDOMINAL PAIN (NON-SPECIFIC), VOMITING/NAUSEA**

**Inclusion criteria for next day clinic:**

- Patient being admitted for abdominal pain or GI symptom control
- CT scan demonstrates no acute etiology of symptoms (e.g., cholecystitis, colitis, diverticulitis), though unrelated pathology is acceptable (e.g., bladder wall thickening, incidental cysts)
- Symptoms due to constipation/large stool burden is acceptable
- Cholelithiasis without cholecystitis (i.e., high suspicion for biliary colic) is acceptable (if not admitted to surgery)
- Still able to drink ~2-4 glasses of water per day, and able to tolerate pills

**Illness-specific exclusion criteria for next day clinic (in addition to general exclusion criteria):**

- CT A/P or H&P concerning for any "red flags" that require surgical evaluation (e.g., SBO, AAA dissection), high risk of decompensation (e.g., toxic megacolon), or any fluid collections concerning for abscesses
- CT with evidence of colitis or diverticulitis (if there is, then refer to that respective algorithm)
- Diarrhea predominant symptoms (if this is the case, refer to the Infectious Colitis & Diarrhea algorithm)
- High suspicion for pancreatitis (if there is, then refer to that algorithm)
- High suspicion for reproductive organ pathology (e.g., ovarian torsion, PID)
- Complete or near-complete PO intolerance of liquids or pills
- WBC > 16,000
- AST or ALT > 3x upper limit of normal; total bilirubin ≥ 4 mg/dL
- Evidence of sepsis by qSOFA (2 out of 3: AMS, SBP≤100, RR≥22)
- QTc ≥ 500 ms

**Treatment in ED:**

1. Order CRP if not already drawn
2. Review or obtain ECG to ensure QTc<500, administer IV ondansetron and/or metoclopramide as usual
    -  Can use 1-2mg lorazepam PO for nausea if refractory to ondansetron/metoclopramide
3. Ensure patient has been adequately rehydrated (e.g., 1-3L crystalloids), based on clinical judgment
4. If there is concern for PUD, then collect h. pylori stool sample (even if patient is on PPI)
5. Standardized guidelines/suggestions for treating <u>abdominal pain</u>
    * In general, <u>use functional pain rather than reported pain (0-10 scale)</u> as more objective measure: if patient can use phone or sleep in ED then they do not need to be admitted. Should be gravely disabled from sx to admit.
    -  Give all patients PO tylenol 1000mg and IV methocarbamol 500-1000mg
    -  If no contraindications and low s/f PUD/gastritis, give ketorolac 30mg IM x1
    -  If opioids are absolutely needed, <u>consider oxycodone 5mg PO</u> (must tolerate PO to qualify for NDC)
6. If imaging reveals massive stool burden, give 30mL milk of magnesia and mineral oil enema q2h x2 enemas
7. If sx responding to above treatment (e.g., able to use phone/sleep; not gravely disabled)  *Schedule Follow-up*
    -  If gravely disabled from pain, please consider another round of treatment and reassess in 1 hr. If still gravely disabled (can't complete ADLs at home), then patient doesn't qualify for NDC-- can admit patient.

*Schedule Follow-up*

1. Confirm that telephone and email (if they have one) are correct
2. Standardized guidelines/suggestions for treating <u>nausea/vomiting</u> at discharge:
    - Orally disintegrating ondansetron 8mg SL q8h prn x30d, +/- lorazepam 1mg PO q6h prn x5d
3. Standardized guidelines/suggestions for treating <u>abdominal pain</u> at discharge:
    - Consider PO methocarbamol, and alternating tylenol and ibuprofen (if no contraindications)
4. Option 1: If clinical judgment suggests additional IV fluids and/or medications (non-opioids) are indicated:
    -  Schedule Next Day Clinic **in-person** appointment within 24 hours

*Dosing must be adjusted for renal impairment unless otherwise specified

5. Option 2: If no further parenteral treatment or lab rechecks are necessary:
    -  Schedule Next Day Clinic **video or phone** appointment in 24-48 hours based on sx severity

**Next day clinic workflow:**

*First follow-up visit (24-48 hours from initial ED visit), telehealth or in-person*

1. Assess for the following red flags: Any hospitalizations at OV or any other hospital, or any unscheduled urgent care or ED visits anywhere, for any cause (record visit dates and length of hospitalization if any). Intractable nausea/vomiting, worsening abdominal pain, fevers/chills/rigors, hematochezia.
    -  If any red flags are present, consult with NDC physician
2. If in-person visit, recheck appropriate labs if needed (e.g., LFTs), provide additional IV treatment (e.g., fluids)
3. If h. pylori was sent in ED, review these results. If positive, then initiate quadruple therapy.
    -  If negative and patient on PPI, inform them it will need to be repeated once PPI can be held x14d
4. If known gallstones and c/f biliary colic, refer to outpatient surgery for evaluation for elective cholecystectomy
5. Assess any patient questions/concerns
    -  If any unexpected concerns, consult with NDC physician
6. Schedule follow-up:
    a. If patient recovering well, no further follow-up needed
    b. If patient requires closer follow-up schedule in ~1-3 days by **video/phone** or **in-person**

*Second follow-up visit (optional; 2-5 days after initial ED visit), telehealth or in-person*

1. Assess for the following red flags: Any hospitalizations at OV or any other hospital, or any unscheduled urgent care or ED visits anywhere, for any cause (record visit dates and length of hospitalization if any). Intractable nausea/vomiting, worsening abdominal pain, fevers/chills/rigors, hematochezia.
    -  If any red flags are present, consult with NDC physician
2. Assess any patient questions/concerns
    -  If any unexpected concerns, consult with NDC physician
3. No further follow-up required. Can schedule regular PCP follow-up

*Dosing must be adjusted for renal impairment unless otherwise specified

### ACUTE KIDNEY INJURY (AKI)

**Inclusion criteria for next day clinic:**

- AKI as defined by: serum Cr >1.5 times baseline OR if unknown baseline then serum Cr >1.5 g/dL
- If there is suspicion for UTI/pyelo, refer to that algorithm instead

**Illness-specific exclusion criteria for next day clinic (in addition to general exclusion criteria):**

- Serum Cr >3.0 times patient baseline
- Patient presents with clinically significant hypervolemia
- Decompensated cirrhosis
- UA: Gross hematuria or microscopic hematuria (>5 RBCs/HPF) with proteinuria
- New proteinuria >300 mg/dL (worsening proteinuria is acceptable)

**Suggested Evaluation/Treatment in ED**

1. Assess UA for gross/microscopic hematuria (>5 RBCs/HPF) with proteinuria OR new proteinuria on dipstick
    a. If any are present then stop algorithm and deliver care as usual
2. Assess BMP for K>6.0, BUN>50, or Cr >3x baseline
    a. If any are present then stop algorithm and deliver care as usual
3. If BMP reveals bicarb <18, then draw VBG
    a. If pH < 7.30 then stop algorithm and deliver care as usual
4. If patient has diabetes or there is concern for BPH/obstructive picture, ensure PVR is obtained in ED
    a. If PVR >250cc, the stop algorithm and deliver care as usual
    b. If PVR is between 100-250cc, then proceed to *Obstructive Algorithm*
5. Consider STAT renal ultrasound if concern for obstruction or unclear etiology of AKI
    a. If new hydronephrosis, then stop algorithm and deliver care as usual

*Likely Pre-renal Etiology:*

1. Administer crystalloids based on clinical judgment
2. Discharge to next day clinic, **in-person** appointment for Cr recheck the following day (~24 hours)

*Likely Post-obstructive Etiology:*

1. Place foley catheter
2. Start tamsulosin 0.4mg qHS if no contraindications.
3. Discharge to next day clinic, **in-person** appointment for the following day (~24 hours)
    a. Prescribe 30d supply of tamsulosin (no refills) at discharge

*Dosing must be adjusted for renal impairment unless otherwise specified

**Treatment in NDC:**

*First follow-up (~24 hours post ED visit), in-person*

1. Obtain BMP and UA immediately upon arrival
    a. If any of the following are present, proceed to Step 2
        - Cr has worsened by >20% since ED visit, new gross/microscopic hematuria (>5 RBCs), new proteinuria >300 mg/dL
    b. If Cr is not improved but has worsened less than 20% since ED visit, proceed to Step 3
    c. If Cr improving, proceed to Step 4. Further fluids can be given per clinical judgment
2. Order STAT renal US (if not already obtained), & page renal who will staff w/ attdg and advise on next steps
    a. In addition to paging renal, place eConsult to nephrology
    b. Stop proceeding down treatment algorithm and defer to renal
3. Schedule a next business day in-person follow-up visit in NDC for repeat creatinine check
    a. Further fluids can be given per clinical judgment
    b. Discharge patient
4. Determine disposition as follows
    a. If patient creatinine improving, plug patient back into PCP in 5-10d for BMP recheck
    b. If patient has foley, in addition to PCP appt place urology eConsult requesting voiding trial in 15-20 days

*For follow-up visits for creatinine recheck, follow the same algorithm as "First follow-up visit"*

*Dosing must be adjusted for renal impairment unless otherwise specified

OV-UCLA Next Day Clinic Inclusion Criteria and Treatment Algorithms
Updated: September 5, 2023

### ASTHMA

**Inclusion criteria for next day clinic:**
- Mild to moderate asthma exacerbations not requiring BiPAP

**Illness-specific exclusion criteria for next day clinic (in addition to general exclusion criteria):**
- Any new O2 requirement in previously naive patient
- Increase in chronic oxygen requirement to > 3L/min nasal canula (target O2 sat ≥90%)
- pH<7.35 on ABG, or pH<7.32 on VBG
- Intubation for asthma in the last 12 months
- Respiratory distress (e.g., marked use of accessory muscles)
- More than 2 prior exacerbations during the past 12 months requiring corticosteroids
- Suspected underlying malignancy on thoracic imaging
- Pneumothorax, or high suspicion for pulmonary embolism
- Acutely decompensated heart failure
- WBC > 16,000/L

**Treatment in ED:**
1. Ensure patient has received 3-4 doses of albuterol/ipratropium nebulizer within the first 1-2 hours, then measure response in peak expiratory flow
   - If post-SABA PEF <60% predicted/personal best, low-threshold to admit patient
   - If post-SABA PEF >60% predicted/personal best, continue down treatment algorithm
2. Prescribe methylprednisone 60mg IV x1 (or equivalent) if no glucocorticoids given yet
3. Prescribe 2g magnesium sulfate IV x1 if there is no renal failure
4. Proceed to *Schedule Follow-up*

*Schedule Follow-up*
1. Confirm that telephone and email (if they have one) are correct and dispense pulse oximeter
2. Schedule **video or telephone** follow-up within 18-24 hours
3. Prescribe inhalers to **quadruple** the home ICS dose as follows (and teach proper inhaler technique):

| If home regimen is this… | …then discharge with this |
|---|---|
| No inhaler or albuterol alone | Budesonide/formoterol (180/4.5) 2 puffs BID x30d |
| Home low dose ICS (e.g., budesonide/formoterol 80/4.5) | Fluticasone/salmeterol (Advair) (500/50) 1 puff BID x14 days (then return to home inhaler) |
| Home moderate dose ICS (e.g., budesonide/formoterol (Symbicort) 160/4.5) | Continue home inhaler + additional budesonide alone (Pulmicort) 180mcg BID x 14 days (then return to home inhaler) |
| Home high dose ICS (e.g., fluticasone/formoterol (Advair) 500/50) | Continue home inhaler + additional budesonide alone (Pulmicort) 360mcg BID x 14 days (the return to home inhaler) |

4. Prescribe additional albuterol rescue inhaler, 2 puffs q4h prn (maximum 12 puffs per day)
5. Prescribe prednisone 40mg PO qday x4 days

*Dosing must be adjusted for renal impairment unless otherwise specified

OV-UCLA Next Day Clinic Inclusion Criteria and Treatment Algorithms
Updated: September 5, 2023

**Next day clinic workflow:**

*First follow-up visit (18-24 hours), telehealth*

1. Assess for the following red flags: worsening chest pain, dyspnea, SOB, cough. Any hospitalizations at OV or any other hospital, or any unscheduled urgent care or ED visits anywhere, for any cause (record visit dates and length of hospitalization if any). Any hemoptysis, palpitations, or lower extremity edema/tenderness (if not previously present), problems with O2 (if discharged with new oxygen).
    - □ If any red flags are present, consult with NDC physician
2. Ask the patient to obtain their ambulatory SPO2 (if not possible then resting SPO2)
    - □ If SPO2 <90%, consult with NDC physician. Otherwise, continue current management.
3. Assess any patient questions/concerns
    - □ If any unexpected concerns, consult with NDC physician
4. Request outpatient pulmonary follow-up
5. Schedule second NDC follow-up visit (in ~48 hours) by **video or telephone**

*Second follow-up visit (72 hours from initial ED visit), telehealth*

1. Follow steps 1-3 of the *First follow-up visit* instructions
2. Ensure patient has pulmonology follow-up scheduled
3. Schedule third follow-up visit (in ~2 days) by **video or telephone**

*Third follow-up visit (~5 days from initial ED visit), telehealth*

1. Follow steps 1-3 of the *First follow-up visit* instructions
2. Instruct patient to de-escalate their higher dose ICS in 9 days (14 days after ED visit) as follows:
    a. Prior home regimen no ICS: budesonide/formoterol 80/4.5 2 puffs BID indefinitely
    b. Prior home ICS low dose: budesonide/formoterol 160/4.5 2 puffs BID indefinitely
    c. Prior home ICS moderate dose: fluticasone/salmeterol 500/50 1 puff BID indefinitely
    d. Prior home ICS high dose: continue high dose ICS and defer to pulm on outpatient follow-up

*Dosing must be adjusted for renal impairment unless otherwise specified

### COPD

**Inclusion criteria for next day clinic:**

- Mild to moderate COPD exacerbations not requiring BiPAP

**Illness-specific exclusion criteria for next day clinic (in addition to general exclusion criteria):**

- New diagnosis of COPD or unclear diagnosis of COPD flare
- New oxygen requirement and unable to set up same-day home O2
- Increase in chronic oxygen requirement to > 3L/min nasal canula (target O2 sat ≥90%)
- pH<7.35 on ABG, or pH<7.32 on VBG
- Prior COPD exacerbation requiring intubation in last 12 months
- Respiratory distress (e.g., marked use of accessory muscles)
- Suspected underlying malignancy on thoracic imaging
- Pneumothorax, or high suspicion for pulmonary embolism
- Acutely decompensated heart failure
- WBC > 16,000/L

**Treatment in ED:**

1. Ensure that albuterol/ipratropium nebulizer has been given q1h x2-4 hours
2. Administer antibiotics if the patient meets 2 of the following: increased dyspnea, sputum volume, or purulence
   - If pseudomonas risk factors: prior pseudomonas in 12 months, abx in prior 2 months, chronic steroids
     
      levofloxacin 750mg IV unless already given
   - If no pseudomonas risk factors:
     
      ceftriaxone 1g IV unless already given
3. Ensure methylprednisone 60mg IV x1 (or equivalent) has been given
4. Administer 2g magnesium sulfate IV x1 if there is no renal failure
5. If new O2 requirement can attempt to arrange same-day home O2, but will likely require admission
6. Proceed to *Schedule Follow-up*

*Schedule Follow-up*

1. Confirm that telephone and email (if they have one) are correct and dispense pulse oximeter
2. Schedule **video or telephone** follow-up within 18-24 hours
3. Prescribe tiotropium/olodaterol (Stiolto) 2 inhalations qday
4. Prescribe rescue ipratropium/albuterol soft mist inhaler (Combivent) or nebs (Duonebs) 1 treatment q4h prn
   a. Prescribe using clinical judgment (e.g., avoid albuterol if history of inducible tachycardia)
5. Prescribe antibiotics as follows:
   a. If pseudomonas risk factors: levofloxacin 750mg PO qday x6 days
      i. Risk factors: Pseudomonas in prior 12 months, antibiotics in prior 3m, glucocorticoid use, etc.
   b. If no pseudomonas risk factors: cefdinir 300mg BID x7 days
6. Prescribe prednisone 40mg PO qday x4 days, no refills.

*Dosing must be adjusted for renal impairment unless otherwise specified

OV-UCLA Next Day Clinic Inclusion Criteria and Treatment Algorithms
Updated: September 5, 2023

**Next day clinic workflow:**

*First follow-up visit (18-24 hours from initial ED visit), telehealth*

1. Assess for the following red flags: worsening chest pain, dyspnea, SOB, cough. Any hospitalizations at OV or any other hospital, or any unscheduled urgent care or ED visits anywhere, for any cause (record visit dates and length of hospitalization if any). Any hemoptysis, palpitations, or lower extremity edema/tenderness (if not previously present), problems with O2 (if discharged with new oxygen).
2. Ask the patient to obtain their ambulatory SPO2 (if not possible then resting SPO2)
    -  If SPO2 >90%, okay to continue current care.
3. Assess any patient questions/concerns
    -  If any unexpected concerns, consult with NDC physician
4. Request outpatient pulmonary follow-up, if appropriate
5. Schedule second follow-up visit (in ~2-3 days) by **video or telephone**

*Second follow-up visit (3-5 days  from initial ED visit), telehealth*

1. Follow steps 1-3 of the *First follow-up visit* instructions
2. Ensure patient has pulmonology or PCP follow-up scheduled
3. No further NDC follow-up required

*Dosing must be adjusted for renal impairment unless otherwise specified

### PULMONARY EMBOLISM

**Inclusion criteria for next day clinic:**

- Lobar PE, segmental PEs, or subsegmental PEs

**Illness-specific exclusion criteria for next day clinic (in addition to general exclusion criteria):**

- Hemodynamically unstable PE (SBP <100 mmHg at time of evaluation, or based on judgment)
- Evidence of pulmonary infarct on imaging
- New oxygen requirement greater than 2L/min nasal cannula (target O2 sat ≥90%)
- Increase in chronic oxygen requirement to > 3L/min nasal cannula (target O2 sat ≥90%)
- Recent history of significant bleeding or risk factors for bleeding (e.g., severe thrombocytopenia)
- PE occurred while on anticoagulation
- More than one lobar PE
- Receiving treatment for pulmonary hypertension (e.g., endothelin receptor antagonist or prostacyclin)
- High sensitivity troponin ≥ 50 ng/L or BNP > 300 pg/mL
- TTE (if obtained) with RV thrombus/clot in motion or increased RV/LV ratio

**ED workflow:**

1. Order troponin and BNP if not already obtained (see exclusion criteria above)
2. Calculate HESTIA score
   - SBP <100 mmHg at time of evaluation (+1)
   - Thrombolysis or embolectomy needed (+1)
   - Active bleeding or high risk of bleeding (+1)
   - PE diagnosed (dx'd) while on anticoagulation (+1)
   - PE dx'd >24 hrs ago & requiring O2 since dx (+1)
   - Severe pain requiring IV pain meds (+1)
   - Medical/social reason for admission (+1)
   - Severe liver impairment (+1)
   - Pregnant (+1)
   - History of HIT (+1)

   Scan for HESTIA score:
3. If HESTIA score ≥1, then admit (CrCl<30 and small O2 requirement okay for outpt). If score = 0 → proceed
4. If new O2 requirement, work with case management to rapidly setup HH O2. If impossible, can admit
5. If creatinine clearance > 30, prescribe rivaroxaban starter pack with no refills (rivaroxaban 15mg po bid x 21 days, then rivaroxaban 20mg po daily x 7 days)
6. If creatinine clearance < 30, prescribe apixaban 10mg BID x7d, then 5mg BID x 21 days

*Schedule Follow-up*

1. Confirm that telephone and email (if they have one) are correct
2. Schedule video or telephone follow-up within 18-24 hours
3. Dispense pulse oximeter

*Dosing must be adjusted for renal impairment unless otherwise specified

**Treatment in NDC:**

*First follow-up visit*

1. Assess for the following red flags: worsening chest pain, dyspnea, SOB, cough, hemoptysis, palpitations, presyncope/syncope, or lower extremity edema/tenderness (if not previously present).
2. If patient is recovering rapidly without complications, no follow-up needed. Refill DOAC for 3 months.
3. If unprovoked PE then refer to PCP to complete hypercoagulable workup
4. If clinical judgment suggests additional follow-up is needed, schedule as appropriate. Otherwise can discharge from NDC.

OV-UCLA Next Day Clinic Inclusion Criteria and Treatment Algorithms
Updated: September 5, 2023

**PANCREATITIS**

**Inclusion criteria for next day clinic:**

- Non-severe acute pancreatitis without necrotizing features
- Still able to drink ~2-4 glasses of water per day, and able to tolerate pills

**Illness-specific exclusion criteria for next day clinic (in addition to general exclusion criteria):**

- Alcoholic pancreatitis (given concern for continued drinking or withdrawal)
- CT A/P suggests parenchymal or peripancreatic necrosis, or fluid collections (including hemorrhage)
- Complete or near-complete PO intolerance of liquids or pills
- WBC > 16,000
- INR>1.4 and/or platelet <50,000
- AST or ALT > 3x upper limit of normal; total bilirubin ≥ 4 mg/dL
- Evidence of sepsis by qSOFA (2 out of 3: AMS, SBP≤100, RR≥22)
- QTc ≥ 500 ms

**Treatment in ED:**

1. Calculate the Harmless Acute Pancreatitis Score:
   Peritonitis on exam (rebound tenderness/guarding), +1 point
   Creatinine ≥ 2 mg/dL, +1 point
   Hematocrit ≥ 43% (male) or 39.6% (female), +1 point
   If 0 points then proceed down algorithm, if ≥1 point then admit
2. If hypovolemic administer ~10cc/kg LR using clinical judgment (if euvolemic, fluid boluses can be harmful)
3. Start maintenance fluids as follows: LR at 1.5cc/kg/hr using clinical judgment
4. Review or obtain ECG to ensure QTc<500, administer IV ondansetron and/or metoclopramide as usual
   Can use 1-2mg lorazepam PO for nausea if refractory to ondansetron/metoclopramide
5. Standardized guidelines/suggestions for treating abdominal pain
   * In general, use functional pain rather than reported pain (0-10 scale) as more objective measure: if patient can use phone or sleep in ED then they may not need to be admitted. Should be gravely disabled from sx to admit.
   Give all patients PO tylenol 1000mg and IV methocarbamol 500-1000mg
   If no contraindications and low s/f PUD/gastritis, give ketorolac 30mg IM x1
   If opioids are needed, consider oxycodone 5mg PO (must tolerate PO to qualify for NDC)
6. If sx responding to above treatment (e.g., able to use phone/sleep; not gravely disabled) *Schedule Follow-up*
   If gravely disabled from pain, please consider another round of treatment and reassess in 1 hr. If still gravely disabled (can't complete ADLs at home), then patient doesn't qualify for NDC-- can admit patient.

*Schedule Follow-up*

1. Confirm that telephone and email (if they have one) are correct
2. Standardized guidelines/suggestions for treating nausea/vomiting at discharge:
   - Orally disintegrating ondansetron 8mg SL q8h prn x30d, +/- lorazepam 1mg PO q6h prn x5d
3. Standardized guidelines/suggestions for treating abdominal pain at discharge:
   - Consider PO methocarbamol, and alternating tylenol and ibuprofen (if no contraindications)
   - Consider oxycodone 5mg q4-6h x7 days if opioids are needed
4. Option 1: If clinical judgment suggests additional IV fluids and/or medications (non-opioids) are indicated:
   Schedule Next Day Clinic **in-person** appointment within 24 hours
5. Option 2: If no further parenteral treatment or lab rechecks are necessary:
   Schedule Next Day Clinic **video or phone** appointment in 24-48 hours based on sx severity

*Dosing must be adjusted for renal impairment unless otherwise specified

**Next day clinic workflow:**

*First follow-up visit (24-48 hours from initial ED visit), telehealth or in-person*

1. Assess for the following red flags: Any hospitalizations at OV or any other hospital, or any unscheduled urgent care or ED visits anywhere, for any cause (record visit dates and length of hospitalization if any). Intractable nausea/vomiting, worsening abdominal pain, fevers/chills/rigors, hematochezia.
2. If in-person visit, recheck appropriate labs if needed (e.g., LFTs), provide additional IV treatment (e.g., fluids)
3. Schedule follow-up:
    a. Option 1: If patient recovering rapidly without complications, no further f/u needed (PCP takes over)
    b. Option 2: If patient requires closer follow-up schedule in ~1-3 days by **video/phone** or **in-person**

*Second follow-up visit (OPTIONAL; 2-5 days after initial ED visit), telehealth or in-person*

1. Assess for the following red flags: Any hospitalizations at OV or any other hospital, or any unscheduled urgent care or ED visits anywhere, for any cause (record visit dates and length of hospitalization if any). Intractable nausea/vomiting, worsening abdominal pain, fevers/chills/rigors, hematochezia.
2. No further NDC follow-up visits required (PCPC can assume care)

### INFECTIOUS COLITIS/DIARRHEA

**Inclusion criteria for next day clinic:**

- CT scan demonstrating non-specific colitis (e.g., colonic wall thickening, fat stranding, mesenteric thickening) and infectious colitis is the leading diagnosis (e.g., fever, hematochezia, profuse diarrhea)
- Still able to drink at least 4 glasses of water per day, and able to tolerate pills

**Illness-specific exclusion criteria for next day clinic (in addition to general exclusion criteria):**

- CT A/P with any “red flags” that require surgical evaluation (e.g., SBO), high risk of decompensation (e.g., toxic megacolon), or any fluid collections concerning for abscesses
- Infectious colitis is the leading diagnosis (if it is, then refer to that treatment algorithm)
- Complete or near-complete PO intolerance of liquids or pills
- WBC > 16,000
- Evidence of sepsis by qSOFA (2 out of 3: AMS, SBP≤100, RR≥22)
- QTc ≥ 500 ms
- Failure of PO antibiotics for current colitis
- Colitis episode within the last 6 months

**Treatment in ED:**

1. Order CRP if not already drawn
2. Send BioFire Stool panel if not already sent (turnaround time 60-90 minutes)
    -  If **only** concern for C. diff colitis, then can order C. diff specific PCR (turnaround time 2 hours)
3. Review or obtain ECG to ensure QTc<500, administer IV ondansetron and/or metoclopramide as usual
    -  Can use 1-2mg lorazepam PO for nausea if refractory to ondansetron/metoclopramide
4. Ensure patient has been adequately rehydrated (e.g., 1-3L crystalloids), based on clinical judgment
5. Standardized guidelines/suggestions for treating abdominal pain
   * In general, use functional pain rather than reported pain (0-10 scale) as more objective measure: if patient can use phone or sleep in ED then they do not need to be admitted. Should be gravely disabled from sx to admit.
    -  Give all patients PO tylenol 1000mg and IV methocarbamol 500-1000mg
    -  If no contraindications and low s/f PUD/gastritis, give ketorolac 30mg IM x1
    -  If opioids are absolutely needed, consider oxycodone 5mg PO (must tolerate PO to qualify for NDC)
6. If sx responding to above treatment (e.g., able to use phone/sleep; not gravely disabled) ⍰ *Schedule Follow-up*
    -  If gravely disabled from pain, please consider another round of treatment and reassess in 1 hr. If still gravely disabled (can’t complete ADLs at home), then patient doesn’t qualify for NDC-- can admit patient.

*Schedule Follow-up*

1. Confirm that telephone and email (if they have one) are correct
2. Prescribe empiric antimicrobial therapy per the table on the following page
    -  **Do not** prescribe empiric antimicrobials for bloody diarrhea if discharging patient
3. Counsel patient on aggressive oral rehydration
4. Standardized guidelines/suggestions for treating nausea/vomiting at discharge:
    - Orally disintegrating ondansetron 8mg SL q8h prn x30d, +/- lorazepam 1mg PO q6h prn x5d
5. Standardized guidelines/suggestions for treating abdominal pain at discharge:
    - Consider PO methocarbamol, and alternating tylenol and ibuprofen (if no contraindications)
6. Option 1: If clinical judgment suggests additional IV fluids and/or medications (non-opioids) are indicated:
    - ⍰ Schedule Next Day Clinic **in-person** appointment within 24 hours
7. Option 2: If no further parenteral treatment or lab rechecks are necessary:
    - ⍰ Schedule Next Day Clinic **video or phone** appointment in 24-48 hours based on sx severity

*Dosing must be adjusted for renal impairment unless otherwise specified

**Treatment in NDC:**

*First follow-up visit (24-48 hours), telehealth or in-person*

1. Assess for the following red flags: Any hospitalizations at OV or any other hospital, or any unscheduled urgent care or ED visits anywhere, for any cause (record visit dates and length of hospitalization if any). Intractable nausea/vomiting, worsening abdominal pain, fevers/chills/rigors, new/worsening hematochezia.
2. If in-person, administer further IV fluids using clinical judgment
3. A reference guide for antimicrobial therapy is on the next page. No need to adjust treatment unless susceptibilities result and the organism is resistant to current treatment.
4. Schedule follow-up:
    a. If patient recovering rapidly without complications, no further NDC f/u required (PCP assumes care)
    b. If patient requires closer follow-up schedule in ~1-3 days by **video/phone** or **in-person**

*Second follow-up visit (OPTIONAL, 2-5 days after initial ED visit), telehealth or in-person*

1. Assess for the following red flags: Any hospitalizations at OV or any other hospital, or any unscheduled urgent care or ED visits anywhere, for any cause (record visit dates and length of hospitalization if any). Intractable nausea/vomiting, worsening abdominal pain, fevers/chills/rigors, new/worsening hematochezia.
2. If in-person, administer further IV fluids using clinical judgment
3. A reference guide for antimicrobial therapy is on the next page. No need to adjust treatment unless susceptibilities result and the organism is resistant to current treatment.
4. No further NDC follow-up needed (PCP can assume care)

OV-UCLA Next Day Clinic Inclusion Criteria and Treatment Algorithms
Updated: September 5, 2023

| Organism: | Suggested Therapy Pending Susceptibilities | Comments |
|---|---|---|
| • Campylobacter (jejuni, coli, and upsaliensis) | Azithromycin 500 mg PO daily x 3 days (alt: ciprofloxacin 500 mg po BID) | Not indicated unless severe (fever, bloody diarrhea), older, immunocompromised, >1 week or recurrent symptoms |
| • Clostridiodies difficile (toxin A/B) | Oral vancomycin 125 mg PO QID x 10 days (alt: metronidazole 500 mg po TID) | |
| • Plesiomonas shigelloides | Ciprofloxacin 500mg PO BID x 3-5 days | |
| • Salmonella | Ciprofloxacin 500mg PO BID x 7 -10 days (14 days if immunocompromised) (alt: azithromycin 500 mg PO daily x7 days, 14 days if immunocompromised) | Nontyphoidal S.enterica tx usually not indicated unless: >50 yo, immunocompromised, vascular grafts, prosthetic joints, hemoglobinopathy, and/or fever with severe diarrhea |
| • Yersinia enterocolitica | TMP-SMX DS PO BID x 5 days (alt: Ciprofloxacin 500 mg po bid x 5 days) | No treatment unless severe or immunocompromised |
| • Vibrio (parahaemolyticus, vulnificus) | Doxycycline 300 mg PO x1 (alt: Azithromycin 1 g po x1) | No treatment unless severe, > 5 days duration |
| • Vibrio cholerae | Doxycycline 300 mg PO x 1 dose (alt: Azithromycin 1 g PO x1) | |
| • Enteroaggregative E. coli (EAEC) | Do not recommend empiric treatment | |
| • Enteropathogenic E. coli (EPEC) | Do not recommend empiric treatment | |
| • Enterotoxigenic E. coli (ETEC) lt (heat labile)/st (heat stable) | Do not recommend empiric treatment | |
| • Shiga-like toxin-producing E. coli (STEC) stx1/stx2 | Do not recommend empiric treatment | **Do NOT** recommend antibiotic treatment-- associated with HUS |
| • E. coli O157 | Do not recommend empiric treatment | **Do NOT** recommend antibiotic treatment-- associated with HUS |

*Dosing must be adjusted for renal impairment unless otherwise specified

| | | |
|---|---|---|
| • Shigella/Enteroinvasive E. coli (EIEC) | Azithromycin 500 mg PO daily x 3 days or Ciprofloxacin 500 mg PO BID x 3 days | If immunocompromised, treat for 7-10 days |
| • Cryptosporidium | Nitazoxanide 500 mg PO BID x 3 days (non-formulary, need TNF request)<br>If immunocompromised (& not HIV), x 2 weeks | Treatment for HIV neg and symptoms >2 weeks. If HIV+ w low CD4, contact ID |
| • Cyclospora cayetanensis | TMP-SMX DS PO BID x 7-10 days | |
| • Entamoeba histolytica | Metronidazole 500mg – 750mg PO TID x 10 days, followed by paromomycin 25-30 mg/kg per day PO in three divided doses x 7 days (for cysts) | |
| • Giardia lamblia | Tinidazole 2gm PO x 1 dose (non-formulary, need TNF request)<br>alt: Metronidazole 250 mg po TID x 5 days | |
| • Adenovirus F40/41 | Do not recommend empiric treatment. | |
| • Astrovirus | Do not recommend empiric treatment | |
| • Norovirus GI/GII | Do not recommend empiric treatment | |
| • Rotavirus A | Do not recommend empiric treatment | |
| • Sapovirus (I, II, IV, and V) | Do not recommend empiric treatment | |

OV-UCLA Next Day Clinic Inclusion Criteria and Treatment Algorithms
Updated: September 5, 2023

### SEGMENTAL COLITIS ASSOCIATED WITH DIVERTICULOSIS (or non-specific colitis)

**Inclusion criteria for next day clinic:**

- CT scan demonstrating non-specific colitis (e.g., colonic wall thickening, fat stranding, mesenteric thickening) and non-infectious colitis is the leading diagnosis
- Still able to drink at least 4 glasses of water per day, and able to tolerate pills

**Exclusion criteria for next day clinic:**

- CT A/P with any “red flags” that require surgical evaluation (e.g., SBO), high risk of decompensation (e.g., toxic megacolon), or any fluid collections concerning for abscesses
- Infectious colitis is the leading diagnosis (if it is, then refer to that treatment algorithm)
- Complete or near-complete PO intolerance of liquids or pills
- WBC > 20,000
- Evidence of sepsis by qSOFA (2 out of 3: AMS, SBP≤100, RR≥22)
- QTc ≥ 500 ms
- Failure of PO antibiotics for current colitis
- Colitis episode within the last 6 months
- Pregnant
- Allograft recipient

**ED workflow:**

1. Order CRP if not already drawn
2. Review or obtain ECG to ensure QTc<500
3. Perform H&P and review labs to ensure no exclusions present, then proceed to *Assignment*
    If any exclusion criteria present, then stop algorithm and deliver care as usual

*Assignment*

1. If paged out Thursday-Sunday *ED Treatment*, if Monday-Wednesday  Admit

*ED Treatment*

1. Administer 1-3L of crystalloids if safe and not already given
2. If anaerobic coverage (e.g., pip-tazo, meropenem) has not been given, administer metronidazole 500mg IV* x1
3. Treat nausea/vomiting with IV ondansetron or metoclopramide provided QTc<500
4. Treat abdominal pain with IV ketorolac (if no contraindications)
    Only give opioids if absolutely needed, and <u>must be given PO</u> (oxycodone 2.5-5mg) to qualify for NDC (If patient has received IV opioids earlier in ED, must demonstrate symptom control with just PO)
5. If symptoms are responding to above treatment (no longer in distress), proceed to *Disposition*
    If patient still in distress from symptoms, given more fluids and alternative anti-emetic and analgesic therapy and reassess in 1 hour. If still in distress then give IV opioids and admit patient.

*Disposition*

1. Confirm that telephone and email (if they have one) are correct
2. Schedule **video or telephone** follow-up in 24-48 hours. Can offer **in-person** visit if you/patient want IV fluids.
3. Prescribe ciprofloxacin 500mg PO q12h and metronidazole 500mg PO q8h x 10 days, no refills
4. Prescribe orally disintegrating ondansetron 8mg SL q8h prn x 30 days, no refills
5. Counsel patient on aggressive oral rehydration therapy

*Dosing must be adjusted for renal impairment unless otherwise specified

OV-UCLA Next Day Clinic Inclusion Criteria and Treatment Algorithms
Updated: September 5, 2023

**Next day clinic workflow:**

*First follow-up visit (24-48 hours), telehealth*

1. Assess for the following red flags: Any hospitalizations at OV or any other hospital, or any unscheduled urgent care or ED visits anywhere, for any cause. Intractable nausea/vomiting, worsening abdominal pain, fevers/chills/rigors, new/worsening hematochezia.
    -  If any red flags are present, consult with NDC physician
2. Assess any patient questions/concerns
    -  If any unexpected concerns, consult with NDC physician
3. Schedule second follow-up visit (in ~5 days) **by video or telephone**

*Second follow-up visit (7 days), telehealth*

1. Assess for the following red flags: Any hospitalizations at OV or any other hospital, or any unscheduled urgent care or ED visits anywhere, for any cause. Intractable nausea/vomiting, worsening abdominal pain, fevers/chills/rigors, new/worsening hematochezia.
    -  If any red flags are present, consult with NDC physician
2. If symptoms not improving but still appropriate for outpatient management, consider adding mesalamine 800mg TID x7 days, no refills.
3. Assess any patient questions/concerns
    -  If any unexpected concerns, consult with NDC physician
4. Schedule second follow-up visit (in ~21 days) **by video or telephone**

*Third follow-up visit (30 days), telehealth*

1. Assess for the following red flags: Any hospitalizations at OV or any other hospital, or any unscheduled urgent care or ED visits anywhere, for any cause. Intractable nausea/vomiting, worsening abdominal pain, fevers/chills/rigors, new/worsening hematochezia.
    -  If any red flags are present, consult with NDC physician
2. Assess any patient questions/concerns
    -  If any unexpected concerns, consult with NDC physician
3. No further follow-up visits required

*Dosing must be adjusted for renal impairment unless otherwise specified

OV-UCLA Next Day Clinic Inclusion Criteria and Treatment Algorithms
Updated: September 5, 2023

### CONGESTIVE HEART FAILURE EXACERBATION (CHF)

**Inclusion criteria for next day clinic:**

- Meets European Society for Cardiology Definition of CHF exacerbation:
  - Typical CHF symptoms (DOE, edema, etc.) **AND**
  - Typical CHF signs (elevated JVP, pulmonary edema, etc.) **AND**
  - Objective evidence of cardiac abnormality (abnormal TTE, elevated BNP, etc.) **AND**
  - There is no other more likely diagnosis to explain the pt’s symptoms (COPD, pneumonia, etc.)
- Low to moderate risk CHF exacerbation as defined below (Emergency HF Risk Grade)
- Documented EF >15% within the past 2 years

**Exclusion criteria for next day clinic:**

- New or increased chronic O2 requirement that is not improving after treatment in ED
- Suspicion for acute coronary syndrome (e.g., presence of typical angina)
- High suspicion for concurrent cardiopulmonary pathology (e.g., COPD flare, PNA)
- If hsTroponin is positive, then 2hr delta troponin ≥20 ng/L (delta <20 ng/L is acceptable)
- Newly discovered CHF
- Coronary revascularization or ICD implantation in the prior 6 months
- Implanted PPM with evidence of PPM dysfunction on ECG (e.g., loss of sinus capture)
- Serum creatinine ≥3.0 mg/dL or over 2x baseline value
- Respiratory distress (e.g., marked use of accessory muscles)
- WBC > 16,000

**ED workflow:**

1. Ensure BNP has been drawn, and that standing weight has been obtained
2. If first hsTroponin positive, ensure a 2 hour troponin is drawn prior to proceeding (can’t increase ≥20 ng/L)
3. Calculate the Emergency Heart Failure Mortality Risk Grade (EHMRG)-- scan code:
4. If low or intermediate risk (<3.5% 7 day mortality) on EHMRG ⍰ *Assignment*
   ⍰ If high risk (≥ 3.5%) then stop algorithm and deliver care as usual (clinical judgment supersedes)

*Assignment*

1. *If paged out Sunday-Wed ⍰ proceed to Treatment in ED, if Thursday-Saturday⍰ Admit to obs*

*ED Treatment*

1. Ensure potassium and magnesium have been repleted to goal 4.5 mEq/L and 2.5 mEq/L, respectively
2. Based on clinical judgment and degree of hypervolemia, consider 2nd dose of IV diuretics (≥2x home dose)
   - Can give as soon as 4 hours after last dose
   - With this second dose, consider giving concurrent PO potassium/magnesium repletion
3. If new O2 requirement, follow separate protocol to arrange same-day home O2 tank/supplies

*Disposition*

1. Confirm that telephone and email (if they have one) are correct

*Dosing must be adjusted for renal impairment unless otherwise specified

2. Schedule follow-up within 24 hours **in-person** for additional IV diuretics and BMP check
3. Prescribe PO diuretics as follows
    a. If exacerbation due to medication noncompliance, can continue home regimen
    b. If patient was compliant with home regimen, double patients home regimen (BID) x7 days, no refills
        i. Also prescribe 20mEq KCl PO BID and magnesium supplementation x7 days
4. Dispense pulse oximeter, instruct patient to weigh themselves without clothes qAM after using restroom

**Next day clinic workflow:**

*First follow-up visit (24 hours from initial ED visit), telehealth or in-person*

1. If in-person, draw BMP + Mg, prescribe IV diuretics and replete electrolytes using clinical judgment
2. Assess for the following red flags: chest pain, dyspnea, SOB, cough, worsening lower extremity edema, palpitations. Any hospitalizations at OV or any other hospital, or any unscheduled urgent care or ED visits anywhere, for any cause (record visit dates and length of hospitalization if any).
    - If any red flags are present, consult with NDC physician
3. Ask the patient to obtain their SPO2
    - If SPO2 <90%, consult with NDC physician. Otherwise continue current management.
4. Obtain the patient's weight from this morning
    - if patient has gained weight, consult with NDC physician
5. Assess any patient questions/concerns
    - If any unexpected concerns, consult with NDC physician
6. Schedule second follow-up visit (in 24-48 hours) by video/phone or in-person if further IV diuresis needed
7. Give patients the following instructions for how to manage their diuretic in between visits:

| Daily Weight Change From Yesterday | Patient Action |
|---|---|
| Greater than 5 lb loss | Halve diuretic and call physician |
| 0-5 lb loss | Continue diuretics without changes |
| 0-2 lb gain | Continue diuretics without changes |
| Greater than 2 lb gain | Double diuretic dose & call physician |

*Second follow-up visit (2-3 days after initial ED visit), telehealth or in-person*

1. Follow steps 1-6 of the *First follow-up visit* instructions
2. Schedule third follow-up visit (~5 days from initial ED visit) **in-person to check labs**

*Third follow-up visit (5 days after initial ED visit), in-person*

1. Order BMP and Magnesium immediately upon arrival
    a. Replete to K/Mg of 4.5/2.5, and adjust outpatient KCl and magnesium prescription as needed
2. Follow steps 2-6 of the *First follow-up visit* instructions
3. If patient was dispensed O2 in the ED, follow protocol to repatriate O2 tanks
4. Decrease patient diuretic dose using clinical judgment to avoid overdiuresis and maintain euvolemia

*Dosing must be adjusted for renal impairment unless otherwise specified

5. No further NDC follow-up needed (PCP and/or cardiologist to assume care)
    a. If patient follows with cardiology, ensure that appropriate follow-up has been requested

OV-UCLA Next Day Clinic Inclusion Criteria and Treatment Algorithms
Updated: September 5, 2023

### PNEUMONIA (COMMUNITY ACQUIRED)

**Inclusion criteria for next day clinic:**

- Moderate-risk (see risk scores below) community acquired pneumonia that would otherwise be admitted
  - Consider for CURB-65 of 1-2 (clinical judgment supersedes scores)

**Illness-specific exclusion criteria for next day clinic (in addition to general exclusion criteria):**

- CURB-65 of 3 or more
- Complicated effusion that will likely require thoracentesis
- Patient has unstable vitals or still actively septic after fluid resuscitation/treatment in ED
- New oxygen requirement and unable to set up same-day home O2
- Increase in chronic oxygen requirement to > 3L/min nasal cannula (target O2 sat ≥90%)
- pH<7.35 on ABG, or pH<7.32 on VBG
- Respiratory distress (e.g., marked use of accessory muscles)
- Suspected underlying malignancy on thoracic imaging
- Pneumothorax, or high suspicion for pulmonary embolism
- Acutely decompensated heart failure

Scan for CURB-65:

**Treatment in ED:**

1. Calculate CURB-65 by scanning the QR code --->
   ⍰ If high-risk (e.g., CURB-65 ≥ 3), consider admission

   ⍰ If low-moderate risk, continue down algo if appropriate
2. If no risk factors for pseudomonas, IDSA (2019) recommends:
   - Ceftriaxone 1g IV and doxycycline 100mg IV while in the ED
   - Discharge w/ doxycycline 100mg q12h x4d AND cefpodoxime 400mg* q12h x4d
   - Patient can get additional doses of IV/IM ceftriaxone q24h in NDC if needed
3. If pseudomonas in respiratory samples in prior 12 months, IDSA (2019) recommends:
   - Obtain sputum cultures
   - Please give IV levaquin 750mg* in ED before discharge (since it's q24h). Can get more IV doses in NDC
   - Discharge w/ levaquin 750mg* qday PO x4 days
4. If new O2 requirement can attempt to arrange same-day home O2, but will likely require admission
5. Proceed to *Schedule Follow-up*

*Schedule Follow-up*

1. Confirm that telephone and email (if they have one) are correct and dispense pulse oximeter
2. Schedule Next Day Clinic in-person appointment within 24 hours for more IV antibiotics
   a. Schedule NDC in-person appointment in 24 hours

*Dosing must be adjusted for renal impairment unless otherwise specified

**Treatment in NDC:**

*First follow-up visit (18-24 hours from initial ED visit), in-person or telehealth*

1. Assess for the following red flags: worsening chest pain, dyspnea, SOB, cough. Any hospitalizations at OV or any other hospital, or any unscheduled urgent care or ED visits anywhere, for any cause (record visit dates and length of hospitalization if any). Any hemoptysis, palpitations, or lower extremity edema/tenderness (if not previously present), problems with O2 (if discharged with new oxygen).
2. Obtain their ambulatory SPO2, okay to continue ambulatory care if sat ≥90%
3. Administer IV antibiotics as follows:
    a. If respiratory cultures obtained in ED, use those to guide therapy
    b. If no culture data: Ceftriaxone 1g IV or Levofloxacin 750mg IV (based on need for pseudomonal coverage)
4. Schedule second follow-up visit in-person in 24 hours for further IV antibiotics

*Second follow-up visit (~2 days from initial ED visit)*

1. Follow steps 1-3 of the *First follow-up visit* instructions
2. No further NDC follow-up required (PCP can assume care)

*Dosing must be adjusted for renal impairment unless otherwise specified
